# Regulation of protein abundance in normal human tissues

**DOI:** 10.1101/2025.01.10.25320181

**Authors:** Huaying Fang, Lihua Jiang, Felipe da Veiga Leprevost, Ruiqi Jian, Joanne Chan, Dafni Glinos, Tuuli Lappalainen, Alexey I. Nesvizhskii, Alexander P. Reiner, GTEx Consortium, Michael P. Snyder, Hua Tang

## Abstract

We report a systematic quantification of 10,841 unique proteins from over 700 GTEx samples, representing five human tissues. Sex, age and genetic factors are associated with variation in protein abundance. In total, 1981 cis-protein quantitative trait loci (cis-pQTL) are identified, of which a majority of protein targets have not been assayed in the recent plasma-based proteogenomic studies. Integrating transcriptomic information from matching tissues delineates concordant as well as discordant expression patterns at RNA and protein levels. Juxtaposition of data from different tissues indicates both shared and tissue-specific genetic architecture that underlie protein abundance. Complementing genomic annotation, RNA-based eQTL studies, as well as the recent establishment of plasma-based proteogenomic characterization, tissue-pQTLs shed light on biology underlying genotype-phenotype association of complex traits and diseases.

## Main

Proteins serve as the fundamental workforce driving cellular activities. Perturbation of the proteome directly impacts biological functions, complex traits, and susceptibility to diseases. Yet factors affecting protein variation in normal human tissues remain a missing piece in the genetic regulation puzzle. How do proteins vary among individuals, with age and between sexes? How do the millions of genetic variants across the human genome affect protein expression in a specific tissue? Answers to these questions can significantly enhance our ability to interpret findings from genome-wide association studies (GWAS), connecting GWAS loci and genes they regulate, as well as the biological processes that they perturb to influence traits or disease risks.

The Gene-Tissue-Expression (GTEx) project was established to chart a map of transcriptional regulation across human tissues. The V8 release of the project has discovered that the RNA expression of 94.7% of all protein-coding genes is associated with proximal genetic variants (cis-eQTL) in at least one of the 49 tissues analyzed^1^. Expanding upon GTEx, the Enhancing GTEx (eGTEx) project sets out to generate additional molecular phenotypes on the same tissues, including telomere length, DNA accessibility, histone modifications, DNA and RNA methylation, and protein quantification^2–5^. The ultimate objective of eGTEx is to build a multi-omics resource that facilitates the elucidation of molecular mechanisms responsible for translating genetic variation to complex phenotypes.

As part of the eGTEx efforts, we report here characterization of biological and genetic effects on inter-individual variation of protein abundance. Complementary to a previous eGTEx study^2^, which surveyed protein variation in a few individuals across 32 tissues, the current study analyzes 144 samples in each of five distinct tissues and reveals widespread variation in protein abundance that is influenced by the age, sex and genetics of the donors (Extended Data Fig. 1). The uniform experimental and analytic approaches applied across tissues allow us to examine tissue-dependent age- and sex-effects on protein variation. Leveraging genomic and transcriptomic data from the corresponding tissue samples, we identify nearly two thousand protein quantitative trait loci (pQTLs), where genetic variants are associated with protein levels. These pQTL loci feature both shared and distinct patterns of regulation among tissues as well as between RNA and protein levels. Integrating pQTL findings with GWAS data, we provide examples where altered protein expression serves as a pivotal link in the molecular cascade connecting genetic variation with complex phenotypes. Complementing the recent population-scale, plasma-based, proteogenomic analyses by substantially increasing the proteome coverage^6–9^ (Fig. S1), this study marks a comprehensive endeavor in simultaneously characterizing the genetics of the human proteome across multiple non-diseased tissues.

### Study design and data acquisition

We used liquid chromatography isobaric tandem mass tag (TMT)-based quantitative mass spectrometry to quantify protein abundance of 720 GTEx samples from five tissues: sigmoid colon, heart left ventricle, liver, lung and thyroid; hereafter these tissues will be abbreviated as colon, heart, liver, lung and thyroid, respectively. Tissue samples analyzed in the current study included donors of both sexes and spanning a broad range of age from 21 to 70 (Table S1). In each mass-spectrometry run, eight samples from a given tissue were analyzed along with two internal reference standards, which represented mixtures of tissue samples (Fig. 1A). Technical replicates of each sample were analyzed in two separate mass-spectrometry runs. Mass spectra were searched against GENCODE v26 translated protein sequence database (GRCh38). Unless otherwise specified, results described here are based on gene-level protein abundance, quantified as the log2 ratio of abundance across isoforms, in a tissue sample relative to an internal reference standard (Methods). Protein abundance was far more correlated between the two replicates of each sample (median Spearman correlation 0.61) than between distinct samples (median Spearman correlation 0.16) of the same tissue. In hierarchical clustering within each tissue, two replicates of the same tissue specimens were almost always (93% of pairs) clustered first (Fig. 1B, Fig. S2). Technical variation was further reduced by incorporating replicates and the second internal reference standards. Two heart specimens were identified as outliers by principal component analysis and were removed from subsequent analyses (Fig. S3). Additional details on protein quantification and normalization procedures are described in Supplementary Information and Fang *et al*^10^.

**Fig 1.**
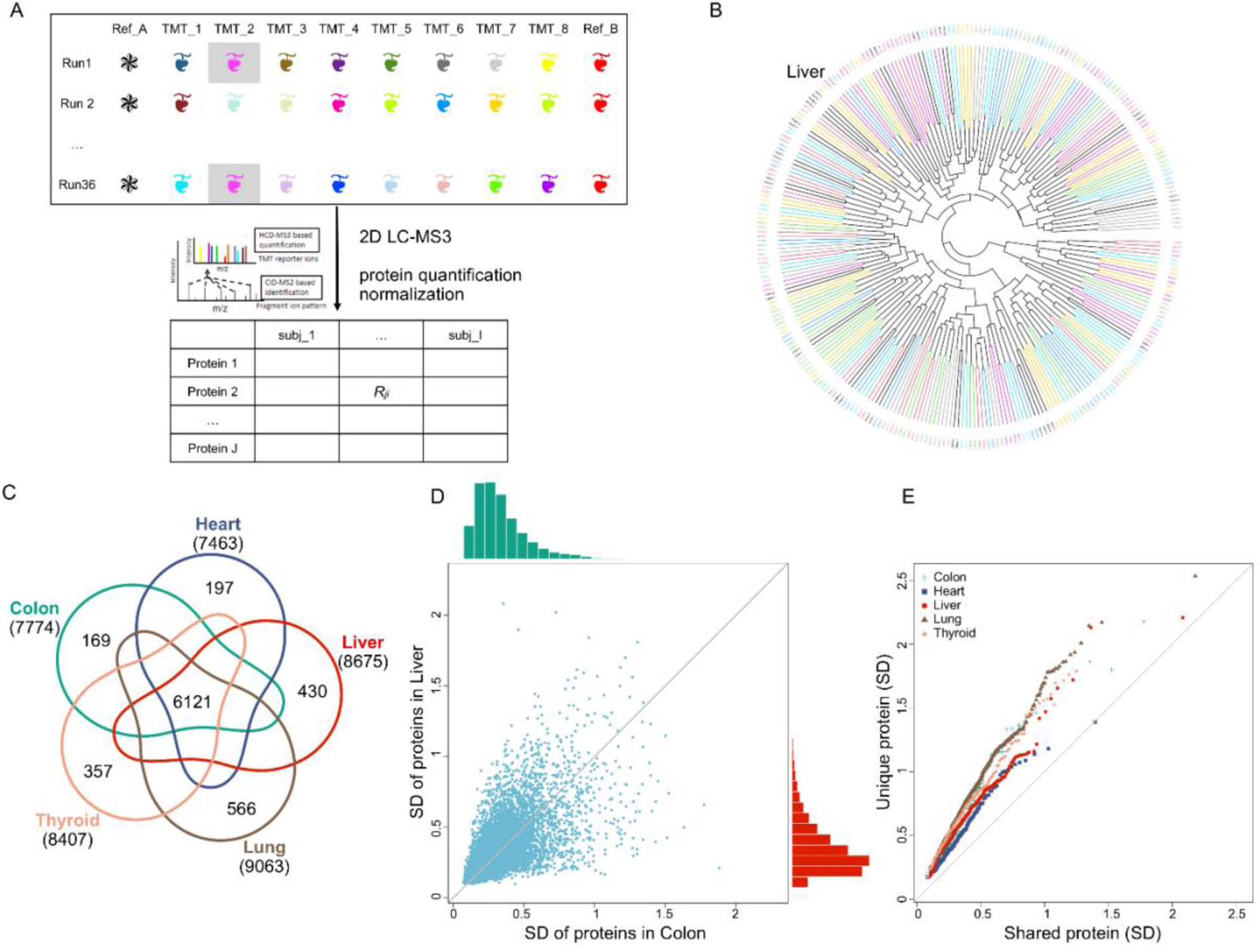
Study overview. (A) Schematic depiction of protein quantification using the tandem mass tag (TMT) mass-spectrometry (MS) platform. Each MS run included eight GTEx tissue samples alongside two reference samples (Ref_A and Ref_B). Every tissue specimen underwent analysis twice, as illustrated by the two shaded squares (Supplementary Information). (B) Hierarchical clustering of protein abundance groups technical replicates of liver tissue specimens. Clustering plots for other tissues are available in Fig. S2. (C) Number of proteins quantified in five tissues. (D) Correlation in protein variability (standard deviation, SD) between colon (x-axis) and liver (y-axis). (E) Quantile-quantile (QQ) plot comparing the variability (SD) of shared proteins (x-axis, defined as proteins present in all five tissues) versus tissue-specific proteins (y-axis).

### Extensive protein variation between individuals

A total of 10,841 unique proteins were quantified in at least one tissue sample, with the majority (N=6121) detected in at least one specimen across all tissues (Fig. 1C, Table S2). Gene ontology (GO) analysis revealed that proteins shared by all tissues were enriched for essential cellular functions, such as transcription and translation. In contrast, proteins unique to a single tissue often engaged in specialized processes (Table S3). For instance, proteins found exclusively in the liver but not in other tissues (N=430) were enriched for cholesterol and sterol homeostasis, while lung-specific proteins (N=566) were associated with axoneme assembly and organization, the core microtubule structure of cilia. Variation of the abundance of a protein across individuals spanned a broad spectrum: whereas the median inter-quartile range (IQR), representing the log ratio between the 75-percentile to the 25-percentile, was 0.40 across five tissues, 5.6% of the proteins displayed an IQR>1, amounting to a two-fold difference. Proteins involved in modulating immune response, such as the regulation of immunoglobin production and complement activation, were enriched among the most variable proteins (Table S3). For proteins found in multiple tissues, inter-individual variation was moderately correlated across tissues: a protein displaying high variability in one tissue tended to exhibit high variability in the other four tissues, with pairwise correlation in standard deviation ranging between 0.63 to 0.75 (Fig. 1D, p<2×10⁻^16^ for each pair). On the other hand, proteins found in only one of the five tissues were, on average, more variable than proteins observed in all five tissues (Fig. 1E), a pattern that remained statistically significant after adjusting for potential confounding factors (p<5.0×10⁻^8^ for each tissue, Supplementary Information, Fig. S6). These observations support a model in which proteins participating in immune responses and tissue-specific functions are subject to stronger dynamic regulation and weaker constraints, resulting in increased inter-individual variation. Notably, 62% proteins (N=6722) quantified in the tissues were not included in the recent plasma-based proteogenomic characterizations, which used targeted detection methods ^7–9^ (Fig. S1).

### Sex and age are associated with protein abundance

Many human complex traits differ between sexes and vary with age. Each tissue features proteins whose abundance are significantly associated with these bio-demographic characteristics of the donors (Fig. 2A, Table S4, Methods). We observed that, in general, the effects of sex on protein abundance exhibited greater heterogeneity across tissues than age effects. This pattern was replicated when comparing the sex- and age-effects in each GTEx tissue with their corresponding impacts on plasma protein abundance^7^ (Extended Data Fig. 2).

**Fig 2.**
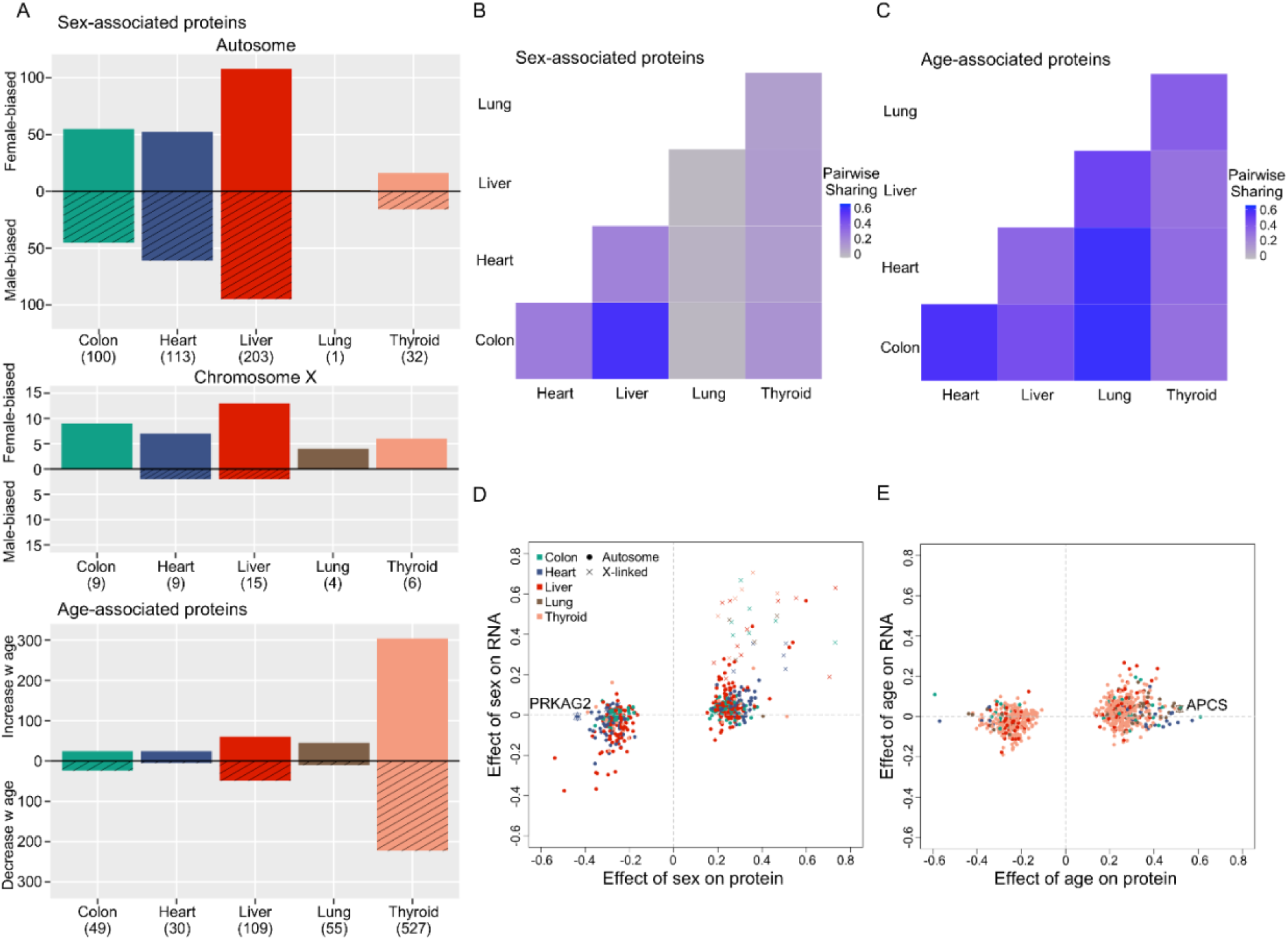
Associations between Protein Variation and Biological Variables. (A) Number of proteins associated with sex and age (at FDR<0.1) and the direction of association. For sex-associated proteins, those encoded by autosome (top) and X-linked (middle) genes are summarized separately. (B) Tissue-sharing of sex-associated proteins. Protein encoded by X-linked genes were excluded. (C) Tissue-sharing of age-associated proteins. (D) Effect size of donor’s sex on protein (x-axis) versus RNA (y-axis) abundance. *PRKAG2* was an example that shows sex-association at protein, but not RNA, level. (E) Effect size of donor’s age on protein (x-axis) versus RNA (y-axis) abundance. APCS was an example that shows age-association at protein, but not RNA, level. (D-E) To depict sex- and age-association on a comparable scale, effect sizes were scaled by sample standard deviation of sex (in D) and age (in E), respectively.

Across the five tissues, 492 protein-sex associations – 221 male-biased and 271 female-biased – were identified, representing 455 unique proteins (at a false discovery rate, FDR<0.1, Fig. 2A). Forty-three associations are located on the X chromosome, a 3.0-fold enrichment relative to the chromosomal distribution of all measured autosomal proteins (Fisher’s exact test p=1.4×10⁻^9^). Whereas the direction of sex-biased protein expression is nearly equal for autosomal genes (51.7% higher in female, p=0.51 using a two-tailed binomial test), 90.7% of the sex-biased protein expression at X-linked genes is in the direction of female-abundant (binomial test p=3.1×10^−8^), suggesting that escape from X-chromosomal inactivation may be a primary driving mechanism. Indeed, 13 of the 24 unique X-linked proteins showing sex-biased protein expression are known to escape X-chromosome inactivation^11^. Among autosomal genes, the effect size of sex on protein abundance exhibited substantial heterogeneity across tissues (Extended Data Fig. 2). An empirical Bayes approach that jointly models all tissues estimated that, on average, 20.9% of the sex-associations are shared between pairs of tissues^12^ (Fig. 2B, Supplementary Information). The pregnancy zone protein (PZP), a secreted protein with female-biased expression, is the only sex-biased protein encoded by an autosomal gene found in more than two of the five GTEx tissues (p=0.12 in heart, p<5×10⁻^4^ in the other tissues). A similar lack of tissue-sharing in sex-differential expression has been observed at the transcriptome level^13^.

Many, but not all, genes with sex-biased protein expression generally displayed a consistent pattern of sex-bias at the RNA level (Spearman correlation of effect size r=0.61, Fig. 2D). A striking example of discordance was observed in a gene located on chromosome 7, *PRKAG2*, which encodes the gamma subunit of AMP-activated protein kinase (AMPK), an energy sensor protein kinase regulating cellular energy metabolism. In heart, the mean PRKAG2 protein abundance is nearly two-fold higher in males relative to females (p<1×10⁻^8^). The male-biased, left ventricular, protein expression has also been reported in mice^14^. In contrast, the *PRKAG2* RNA expression shows no significant difference between sexes in left ventricle despite the larger sample size (p=0.77), nor was this gene reported in the top 500 genes exhibiting sex-biased RNA expression in any of the 44 GTEx tissues^13^.

The 770 protein-tissue pairs, which varied significantly with donor’s age (FDR<0.1), are distributed across the genome. Of the 718 unique proteins, 527 were found in thyroid and 109 were found in liver, while much fewer were observed in colon, heart, or lung (Fig. 2A, Table S4). Age-associated gene expression variation is generally consistent at the RNA and protein levels, although the agreement is weaker compared to that for sex-biased expression (Spearman correlation in effect size r=0.4, Fig. 2E). An example highlighting the discordance at the RNA and protein levels was found in *APCS*, whose protein level exhibited the strongest positive association with age in colon, heart, lung and thyroid (p<1×10⁻^7^ in each tissue, Fig. 2F), and has been reported as one of the three proteins most strongly associated with age in an independent proteomic analysis in skeletal muscle^15^. *APCS* encodes serum amyloid P component (SAP), a component in humoral innate immunity and a constituent of all human fibrillar amyloid deposits. Higher SAP level has been associated with amyloidosis and Alzheimer’s disease^16,17^. RNA expression of *APCS* is measured in GTEx liver and lung, but shows no dependency on age in either tissue.

In contrast to the pervasive tissue-heterogeneity in sex-biased protein expression pattern, 39.3% of age-associated protein expression were shared between pairs of tissues (Fig. 2C, Extended Data Fig. 2). Gene sets related to humoral immune response are significantly enriched for proteins whose abundance increased with age, whereas those involved in the regulation of RNA splicing, spliceome, and oxidative phosphorylation are enriched for proteins whose abundance decreases with age (Table S3). Of note, age-associated alterations in spliceosome protein expression were found in liver, lung and thyroid in our study, and has been reported in human muscle^15^. These observations support the roles of inflammation, oxidative stress and mitochondrial dysfunction as hallmarks of aging, as well as a putative mechanism in age-related chronic diseases, in which age-related deregulation of mRNA splicing triggers cells to enter senescence^18^.

### Protein-RNA and protein-protein associations

The proportion of protein variation between individuals that can be recapitulated by RNA variation has important implications for deciphering the mechanisms underlying genotype-phenotype associations. With the exceptions of a few studies^19^, gene-level correlation between RNA and protein across individuals has not been systematically assessed across non-disease human tissues. The relatively large number of samples assayed in each tissue afforded the opportunity to fill this gap (Table S2). Unsurprisingly, over 74% of all genes displayed positive correlations between RNA and protein abundance within each tissue examined (binomial test p<1×10⁻^16^ for all tissues). However, the magnitude of the correlation was generally low, with a median correlation of 0.092 across all five tissues examined and regardless of covariate adjustments (Fig. 3A, Fig. S7). We hypothesize that the discordance reflects, in part, post-transcriptional regulatory mechanisms that give rise to differential dynamics between synthesis and turnover at the RNA and protein levels. Genetic factors that impact protein abundance without apparent altered RNA expression are discussed in the next section. We reasoned that genes, whose functions featured temporal variation, might be under synchronized regulation and thus display higher protein-RNA correlation across individuals^20^. One such group of genes, exhibiting circadian rhythm at the transcriptomic level, have recently been identified in hepatocytes and have functions related to drug absorption, distribution, metabolism, and excretion (ADME)^21^. Protein-RNA correlations of ADME proteins were indeed significantly higher than non-ADME genes (median correlation r=0.26, two-tailed Wilcoxon test p<1×10⁻^16^, Fig. 3B). A member of this group, *GSTM3*, was among proteins with the highest RNA-protein correlation in all five tissues (Table S2). The strong correlation was partially explained by cis-genetic variations (tagged by rs4970777), which acted both as a cis-eQTL and a cis-pQTL (Fig. 3B inset). RNA and protein remained correlated within each genotype, indicating additional regulatory effects of RNA that were propagated at the protein level. In contrast, stably transcribed genes, such as ribosome and proteasome complexes, featured significantly lower RNA-protein correlations (median correlation 0.05, p<1×10⁻^9^).

**Fig 3.**
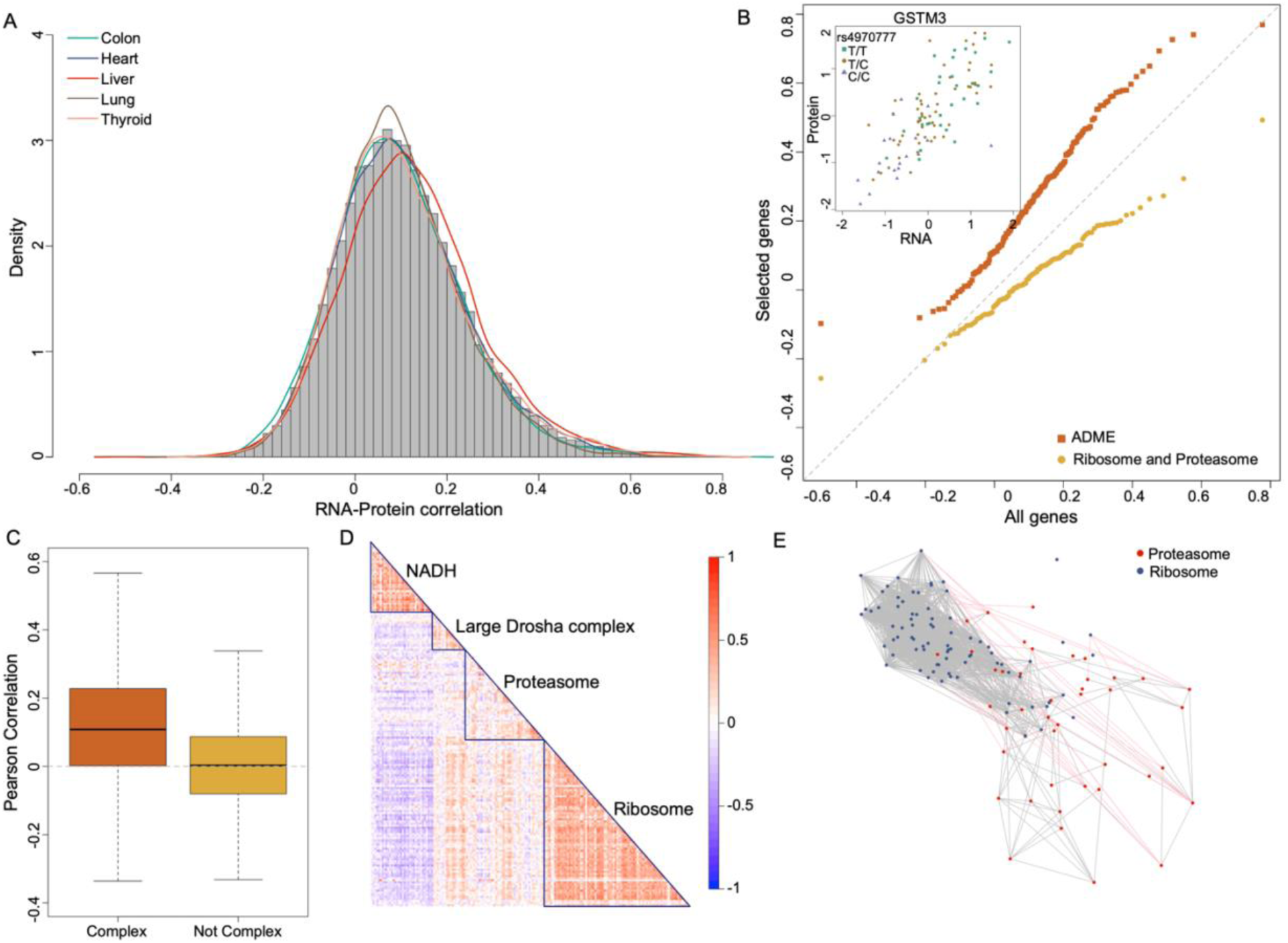
Protein-RNA and Protein-Protein Correlations. (A) Protein-RNA correlation for each gene across individuals. Pearson correlation was computed within each tissue and combined in the histogram. Density curves depicted the distributions of the correlation for each tissue. (B) QQ-plot comparing the distributions of liver RNA-protein correlation of all genes (x-axis) to ADME genes (absorption, distribution, metabolism, and excretion, orange), and to genes in ribosome and proteasome complexes (gold). Inset: One of the ADME genes, *GSTM3*, featured among the strongest RNA-protein correlation in liver (r=0.77). (C) Protein-protein correlation across individuals between gene-pairs in known protein complexes (left) or those not in known protein complexes (right). Boxplots show interquartile range (IQR) and median, with whiskers extending to ±1.5 × IQR. (D) Heatmap of pair-wise protein correlation among genes defining four large complexes in colon. (E) Network representing covariation of proteins in two complexes in colon: ribosome (blue nodes) and proteasome (red nodes). Two nodes are connected if and only if their correlation exceeds 1%-quantile among all pairwise protein correlations. Edges in pink indicate covariation between proteins in different complexes.

Many proteins function as subunits in protein complexes and may be under coordinated regulation of synthesis, maintenance and degradation. We found that proteins in a complex display, on average, stronger pairwise correlation compared to proteins not in a complex (p<1×10⁻^4^, Fig. 3C, Supplementary Information); Fig. 3D displays the co-variation pattern among four large protein complexes in colon. Furthermore, Fig. 3E illustrates the co-expression patterns within and between two functionally related protein complexes in colon: ribosome, the central machinery for protein synthesis, and proteasome, a major component in protein degradation. The co-expression patterns were statistically significant for proteins within each complex (p<1×10⁻^4^) as well as for proteins across the two complexes (p=0.0082, Supplementary Information). These results, along with the trans-pQTL results presented below, suggest that both physical interaction and related functions can play important roles in orchestrating protein abundance.

### Mapping proximal protein quantitative trait loci (cis-pQTL)

Treating relative protein abundances as molecular phenotypes, we searched for cis-associated variants located within 1 MB of the protein-coding region (Methods). Across five tissues, we identified a total of 1,981 cis-pQTLs at FDR<0.1. Following the eQTL nomenclature, we refer to the index SNP at a pQTL locus, the variant with the lowest p-value, as the pSNP, and the associated protein phenotype as the pGene. Given the limited power of fine mapping due to modest sample sizes^22^ (Supplementary Information), subsequent analyses primarily use pSNPs as proxies for the drivers of the pQTLs.

While cis-pQTLs were analyzed within a large window surrounding the coding region, the distance between the pSNP and the transcription start site (TSS) of the pGene is typically much shorter, with a median distance of 32 KB (Extended Data Fig. 3). Among the 1,012 pSNPs located outside the transcribed region of the pGene, 28% were associated with the nearest gene (Fig. 4A). Even in gene-rich regions where a pSNP had more than 50 potential gene targets, 45% of the associated proteins were encoded by one of the three physically closest genes. Conversely, among the 579 pSNPs with more than 20 nearby genes, 150 had target proteins located farther than 10 genes away. As pSNPs were defined based on p-value alone, the true causal variants of the cis-pQTLs are likely situated even closer to the pGenes. These findings suggest that physical proximity to a gene serves as an informative, albeit imperfect, indicator of a pSNP’s functional target.

**Fig 4.**
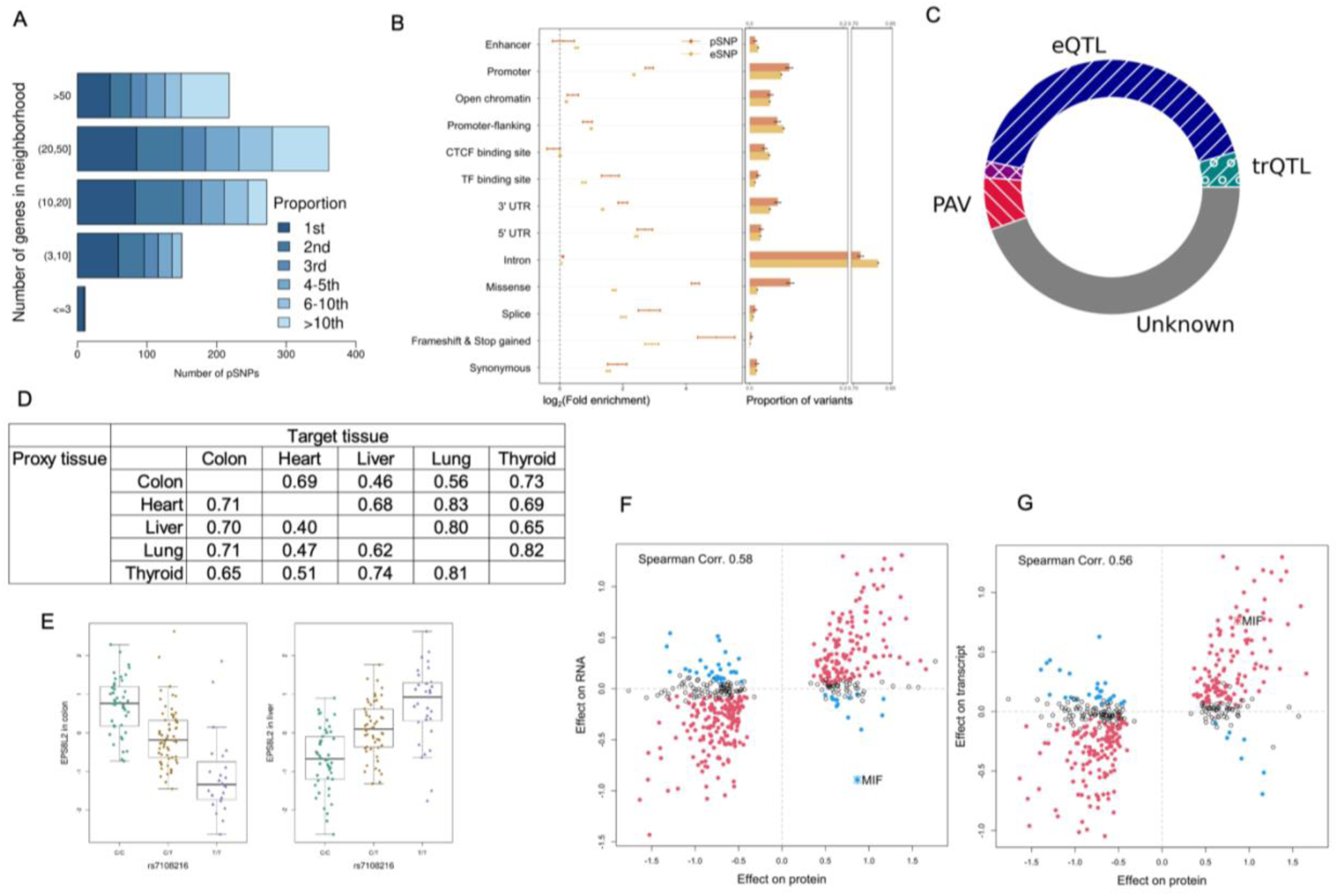
Protein quantitative trait loci (pQTL). (A) Proximity between pSNPs and corresponding pGenes. pQTLs were grouped based on the number of potential pGenes located within the *cis*-neighborhood (1MB) of pSNPs. Colored segments within each group indicate the proximity ranks of the pGene relative to the pSNP. The distribution of physical distance between pSNPs and pGenes is shown in Extended Data Fig. 3. (B) Genomic context of index SNPs at pQTLs (pSNPs, red) and eQTLs (eSNPs, blue). Proportions were normalized to all pSNPs (respectively, eSNPs) within the considered genomic annotations. Enrichment was computed relative to all SNPs tested for pQTLs (respectively, eQTLs). In the forest plot (left), each point represents the point estimate of the log2(Fold enrichment), with the horizontal segment indicating ±1 standard error. The center of each bar plot represents the proportions of variants, with the error bar denoting the standard error. (C) Proportion of pQTLs putatively attributable to molecular mechanisms. The 1981 pQTLs were grouped into three putative mechanisms: protein altering variant (PAV), eQTL, and transcript-QTL (trQTL). trQTL-evidence was only sought when a pQTL was not supported by eQTL. pQTLs supported by both PAV and eQTL/trQTL were denoted in an overlapping section (purple). If none of these mechanisms were supported, the pQTL was assigned to group Unknown (grey, n=883). (D) Proportions of pQTLs identified in one tissue (target) with supporting evidence in another tissue (proxy). (E) EPS8L2 illustrated an example of pQTL with opposite allelic effects observed in colon and liver. Boxplots show interquartile range (IQR) and median, with whiskers extending to ±1.5 × IQR. (F, G) Consistency of allelic effects at protein (x-axis) and RNA (y-axis) levels for pQTLs discovered in liver. (F) RNA was measured by gene-level RNA abundance. (G) RNA represented the RNA-transcript corresponding to protein-coding isoform. pQTLs whose protein-coding isoform was not unique or was not quantified were excluded. As an example, MIF, is high-lighted, showing consistent pQTL and trQTL allelic effects but opposite eQTL allelic effects.

### Transcriptomic-level support of pQTL

Juxtaposing QTLs at the RNA and protein levels offers opportunities to delineate the cascade of regulatory mechanisms across molecular phenotypes.

As expected, cis-eQTLs, which alter gene expression at the RNA level, can have downstream effects on protein abundance. However, significant mediation by RNA expression could only be assigned to 9% (179) of all pQTLs^23^ (FDR<0.1, Supplementary Information, Fig. S12). Using a less stringent criterion of co-occurrence, we defined a pQTL as *eQTL-supported* if the pSNP was significantly associated with RNA abundance in the matching tissue and exhibited the same direction of allelic effects at both the RNA and protein levels. This approach classified 879 (45%) eQTL-supported pQTLs (Fig. 4C). As expected, these eQTL-supported pGenes displayed substantially elevated correlations between RNA and protein abundance compared to both pGenes not supported by eQTLs and genes without a cis-pQTL (Supplementary Information, Fig. S15).

Alternative transcription and alternative splicing of mRNA can generate transcript isoforms, of which only some are translated^24^. Genetic variants that alter transcript structure or isoform ratios represent an important regulatory mechanism for protein expression. Therefore, we examined the relationship between the pSNPs and the abundance of protein-coding RNA transcripts. This analysis identified transcript-level QTLs (trQTLs) as potential drivers for an additional 88 pQTLs, supported by both significant association and consistent direction of allelic effects (Fig. 4C, Supplementary Information). Among these trQTL-supported pQTLs, 44 pQTLs overlapped with splicing QTLs (sQTLs)^25^; the remainder suggested alternative transcriptional mechanisms or reflected more precise RNA quantification. An example of the latter was illustrated by a trQTL-supported pQTL of *MIF* in the liver, where the protein-increasing allele of the pSNP was significantly associated with decreased gene-level RNA abundance (Extended Data Fig. 4, Supplementary Information). The discordant allelic effects were likely due to a partially overlapping antisense RNA (*MIF-AS1*, ENSG00000218537), which was accounted for in transcript quantification but not in gene-level RNA quantification. This complexity underscores the value of long-read sequencing data and improved transcript-level quantification algorithms.

In total, 967 of the 1,981 (49%) pQTLs were supported at the RNA level either by gene-level or transcript-level QTLs, leaving over half of the pQTLs not supported by cis-transcriptomic regulatory evidence, despite the larger sample sizes and higher power of the eQTL analysis compared to pQTL analysis (eQTL sample sizes: colon 318, heart 386, liver 208, lung 515, and thyroid 574). These results, combined with the modest RNA-protein correlation, highlights the important roles of non-overlapping regulatory components affecting protein and RNA expression.

### Genomic context of cis-pQTL associations

Integrating pQTL with functional annotation of the genome enables us to explore the molecular mechanisms that underlying protein variation. Cis-pSNPs are enriched in functional genomic elements. Given the co-occurrence of cis-eQTL and cis-pQTL, it is not surprising that functional elements enriched for eSNPs, such as open chromatin regions, transcription factor (TF) binding sites and 5’-UTR, were likewise enriched among pSNPs. More surprisingly, promoters and the 3’-UTR, showed stronger enrichment for pSNPs compared to eSNPs. These regions are known to influence translation through mechanisms such as the stability and subcellular localization of mRNAs^26,27^.

Protein-altering variants, including missense variants, splice donor and acceptor sites, and variants leading to frameshifts or changes in stop codons, are expected to change protein levels (Fig. 4C). When a pSNP is a protein-altering variant (PAV), the pQTL may reflect changes in protein abundance due to disrupted translation, stability, or degradation. Alternatively, it may arise from the presence of non-reference peptide variants, analogous to the epitope effect in affinity-based proteomics^28^. Of the 172 pQTLs where the pSNPs were PAVs, 87% of non-reference alleles were associated with decreased protein abundance (Extended Data Fig. 5, Supplementary Information). In comparison, 64% of these pQTLs were associated with decreased RNA abundance, suggesting that the directional bias was not solely due to the under-detection of non-reference peptide variants. In some cases, excluding peptides carrying PAVs can help distinguish pQTLs that represent actual changes in protein abundance from those reflecting biased detection of non-reference peptide variants (an investigation of the pQTL at *NAT2* is provided in Supplementary Information as an example, Extended Data Fig. 8). However, this approach may increase protein quantification noise and reduce power for mapping pQTLs, particularly in proteins with multiple PAVs^29^.

### Tissue-sharing and specificity of pQTLs

The 1,981 cis-pQTLs represent 1,237 unique pGenes: 852 genes are statistically significant in exactly one tissue, while 58 genes are significant across all five tissues (Table S5). To estimate the degree of tissue-sharing, we employed two complementary approaches, considering that discordance could be partly due to moderate statistical power (Supplementary Information). In pairwise tissue comparisons—treating one tissue as a discovery sample and another as a replication sample— between 40% and 83% of the pQTLs identified in one tissue were likely shared in another (Fig. 4D, Fig. S11). Consistent with this analysis, a Bayesian meta-analysis that combined all tissue-specific pQTL mapping results suggested that 45% of the pQTLs are shared by all five tissues, while fewer than 10% are tissue-specific (Extended Data Fig. 6A).

In most cases, the directions of allelic effects were consistent across tissues: the pSNP allele associated with increased abundance in one tissue is likely to be associated with increased abundance in another (Extended Data Fig. 6B). However, we also observed pQTLs with discordant allelic effects across tissues. A prominent example is the actin-binding protein EPS8L2: the T-allele of rs7108216 is associated with increased protein abundance in the liver (p=3.4×10⁻^14^) but decreased abundance in the colon (p=5.7×10⁻^22^) (Fig. 4E). This discordance in pQTL effects is supported at the RNA level, showing significant and consistent directional effects in the corresponding tissues. The observed tissue-specific opposite effects may arise from a single regulatory variant—either rs7108216 itself or one in linkage disequilibrium (LD) with it—that confers tissue-specific effects. Alternatively, rs7108216 may tag multiple regulatory variants, each active in different tissues. Regardless of the precise identity of the causal variant(s), these discordant allelic effects underscore the tissue-dependent component of the gene regulatory landscape. Taken together, these results suggest that cis-pQTLs identified in one tissue can serve as useful proxies for predicting genetic effects on protein abundance in other tissues. However, for specific loci of interest, such predictions must be validated in the relevant tissue or cell types.

Given the substantial overlap in cis-pQTLs across tissues, we sought validation in two independent plasma proteogenomic studies, which employed complementary proteome platform (SOMAscan and Olink arrays) and represented much larger sample sizes for pQTL discovery^8,9^. Because a substantial fraction of proteins quantified in GTEx tissues were not assayed in the plasma studies (Fig. S1), 326 and 683 unique pSNP-pGene pairs identified in GTEx could be looked up in the two plasma datasets, respectively. Despite the technical differences between the proteomic platforms and the biological differences between plasma and GTEx tissues, the allelic effects of the pSNPs were highly correlated in plasma and GTEx tissues with r=0.54 (p=3.5×10^− 27^) and 0.54 (p=5.5×10^−56^), in the two replication datasets, respectively. The replication rate, quantified by the estimated non-null proportion (*π*_1_), were 0.55 and 0.67 in the two datasets, respectively (Extended Data Fig. 7, Fig. S9). Loci that show substantial discrepancy may arise from tissue difference, platform difference or stochastic errors.

### Trans-pQTL

To identify trans-pQTLs, we performed a genome-wide association study (GWAS) treating each protein abundance in each tissue as a quantitative trait (Methods). Across five tissues, 30 trans SNP-protein associations were identified (p<1.96×10^−10^, Table S5). Fourteen of the trans-pSNPs, showed suggestive association with a protein in *cis* (at p<0.01) in the same tissue. Furthermore, for six of these 14 SNPs, the cis- and trans-pQTL targets may interact through protein-protein interaction, indicated by a STRING score greater than 900 (95-percentile among the 11 million pairs of protein-protein scores in STRING-db^30^). Some protein interactions represent subunits in known protein complexes. As an example, a cis-pSNP of IDH3A on chr15 is identified as a trans-pSNP for both IDH3B (chr20) and IDH3G (chrX) in heart. Proteins encoded by *IDH3A*, *IDH3B* and *IDH3G* constitute the subunits (α, β and γ respectively) of the mitochondrial NAD-dependent isocitrate dehydrogenase (IDH3-NAD+). X-ray crystallography has revealed that this protein complex consists of two heterotetramers (α_2_βγ), each of which is, in turn, formed by two heterodimers, αβ and αγ^31^. Therefore, the steady-state abundance of each of the subunits is not only determined by its own synthesis and degradation, but also likely constrained by the stoichiometry ratio between subunits. Other cis- and trans-protein pairs do not form a physical complex but participate in a common pathway or cellular structure. For example, the genotype of rs2076295 was associated with two desmosomal proteins in lung: PKP3 in trans and DSP in *cis*. Of note, rs2076295 was not significantly associated with the RNA level of PKP3 (uncorrected p=0.22, Fig. S9C), indicating that this trans-pQTL was likely not mediated through PKP3 RNA-expression. Together, these results support a model in which cis-genetic regulation of gene expression perturbs distal protein abundance that leads to coregulation of proteins.

In sharp contrast with cis-pQTL, but consistent with an observation in trans-eQTL analyses^1^, trans-pQTLs exhibited limited sharing across tissues: 28 of the 30 SNP-protein associations were observed in exactly one tissue. Of the significant *trans* SNP-protein pairs tested in an independent plasma study^9^, none was replicated after the Bonferroni correction of multiple testing (at p<0.05/7) despite of the much larger sample size. These findings suggest that cell-type composition and tissue-specific factors may play a greater role in the trans-regulation of protein expression than they do in cis-regulation.

### pQTLs provide insights on complex traits associations

Of 1827 unique cis-pSNPs identified in the current study, 603 were significantly associated with one or more phenotypes in the GWAS catalog or phenoscanner (p≤5×10^−8^). Examples include rs10774671, whose alternative allele (A) is associated with a decrease in OAS1, an interferon-inducible antiviral protein (pQTL p=1.12×10^−23^ in liver). This SNP has been previously associated with an increased risk for severe COVID-19 and hospitalization and influences circulating OAS1 protein levels through allele-specific regulation of splicing and nonsense-mediated decay^32,33^. Most individuals of European origin carry the alternative A allele. The ancestral G allele is of Neanderthal origin and is the major allele in African populations. The G allele is associated with production of the p46 OAS1 isoform, which appears to possess greater activity against certain RNA viruses than the A allele-associated p42 isoform^34,35^. Although the co-occurrence of a cis-pQTL and a complex trait GWAS association does not define mechanistic link, below we show examples that demonstrate the type of information pQTLs can add to existing functional genomic data.

To date, exonic variants – including variants in the protein-coding region, 3’- and 5’- UTR as well as splice donor and receptor sites– are commonly linked to the genes in which they reside. However, an exonic variant can act as a cis-regulatory variant on a neighboring gene. Ignoring this possibility can lead to erroneous functional interpretation of GWAS SNPs. A well-known example of this type of variant is rs12740374, associated with low-density lipoprotein cholesterol (LDLC) and CAD. This variant resides in the 3’-UTR of gene *CELR* but also regulates liver gene expression of *SORT1* at both RNA (p=3.1×10^−54^) and protein (p=7.09×10^−20^) levels. However, under an assumption that an “exonic” variants are more likely to impact phenotypes than do non-coding variants regulating transcription, *CELR* would be erroneously favored as the gene target^36^. We identified 67 similar instances, in which a SNP was annotated as exonic for one protein-coding transcript and was found associated with the protein abundance of a different gene in *cis* (Table S6). One such variant, rs10117795, represents a GWAS locus of serum concentration of dipeptidyl peptidase 2 levels^37^. A missense variant in *MAN1B1*, rs10117795 was also the pSNP associated with *DPP7* (p=2.169×10^−19^ in liver). In this case, *DPP7* (ENSG00000176978, also called *DPP2*) is likely the underlying gene, as it encodes dipeptidyl peptidase 2 itself. At the RNA-level, rs10117795 was cis-associated with the expression of *MAN1B1*, *DPP7* and several other genes; hence, the target gene could not be disambiguated based on eQTL evidence alone. In contrast, rs10117795 was not associated with *MAN1B1* at the protein level across the four tissues, in which cis-pQTL was tested. As a second example, SNP rs11191421 is a GWAS index SNP for serum total testosterone level^38^. The variant is located in the 3’-UTR of *BORCS7* gene and associated with protein levels of Arsenite Methyltransferase (*AS3MT*) in colon (p=3.74×10^−5^), heart (p=8.955×10^−6^) and thyroid (p=1.388×10^−4^). *AS3MT* protein catalyzes the metabolism of arsenic, which has, in turn, been shown to cause male reproductive damage and reduced serum testosterone levels ^39,40^. Thus, the pQTL finding generates a testable mechanistic model, in which rs11191421 *GG*-genotype leads to reduced serum total testosterone level via decreased *AS3MT* and impaired arsenic metabolism.

Protein targets at pQTLs can also identify genes and functions that underlie GWAS sites, for which transcriptomic level evidence is absent. At chromosome 17*q*, a liver pQTL of *ABCA8* overlapped with a GWAS locus associated with plasma high-density lipoprotein cholesterol (HDLC) level^41^ (Fig. 5A-C). Fine-mapping constructed the credible set for the causal variant of the pQTL, which consisted of six variants, including the pQTL index SNP, rs1373068, and the HDLC-lead SNP rs4148005, both of which are located in the intron of ABCA8 and were in high LD with each other (r^2^=0.93). RNA analyses did not inform the functional impact of these genetic variants, as neither variant was identified as an eQTL, trQTL or sQTL in a relevant tissue, nor were they implicated in the larger plasma-based eQTLGen Study^42^. Examining transcript-level expression in liver, we did not find an association between the rs4148005 genotype and the protein-producing transcript (ENST00000430352) (p=0.20). Furthermore, although the HDLC-lead SNP is located within 500kb from the transcription start site of eight protein coding genes and three lincRNAs, it was only associated with a functionally uncharacterized lincRNA transcript, RP11-118B18.2 (p=1.1×10^−7^ in liver). Thus, transcriptomic analysis does not inform function underlying this HDLC locus. The pQTL finding, in contrast, postulates a testable molecular cascade in which the HDLC-increasing effect of the rs4148005-G allele is mediated through elevating ABCA8 protein abundance. *ABCA8* is a member of the superfamily of ATP-binding cassette (ABC) transporters. Indeed, rare pathogenic variants of *ABCA8* have been identified among individuals with low HDLC^43^. Furthermore, hepatic overexpression of human ABCA8 in wild-type mice results in increased plasma HDLC, while *in vitro* studies have suggested its role in cholesterol efflux^44^.

**Fig 5.**
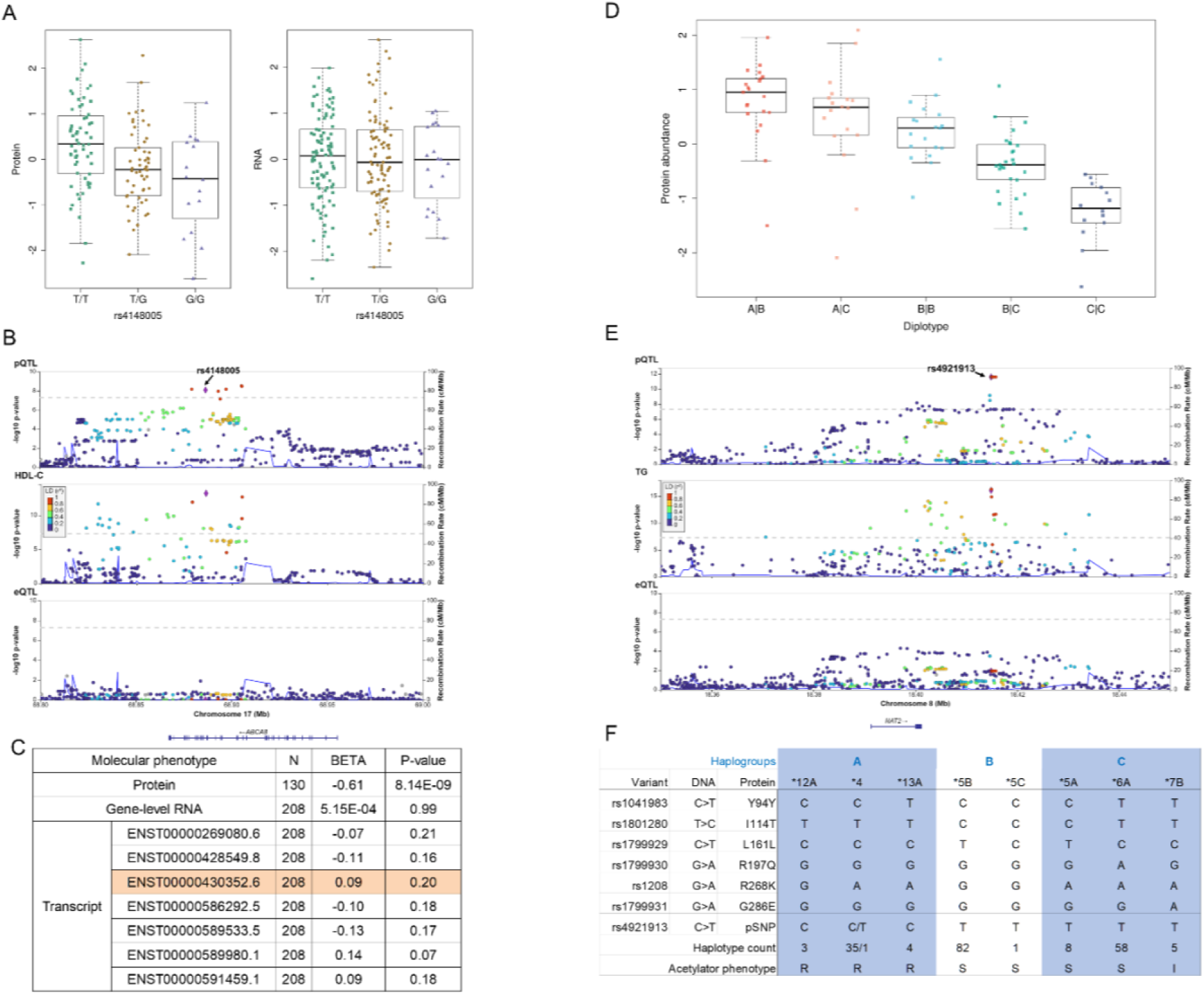
Protein Quantitative Trait Loci (pQTL) Informing Complex Trait Associations. (A-C): pQTL analysis suggests *ABCA8* as a putative link underlying a HDL-C (High-Density Lipoprotein Cholesterol) locus on Chr 17, identified through genome-wide association studies (GWAS). (A) Variant rs4148005 shows association with ABCA8 protein levels but not with RNA levels. (B) The HDL GWAS (middle) index SNP, rs4148005, coincides with the ABCA8 pQTL in liver (top) but lacks association with RNA levels in the region. (C) Summary of associations between the rs4148005 genotype and the abundance of the *ABCA8* protein, RNA and transcript. The protein-coding transcript is highlighted. (D-F): pQTL analysis suggests *NAT2* as a putative link underlying a triglyceride (TG) GWAS locus on chromosome 8. (D) *NAT2* diplotype correlates with protein abundance in the liver. (E) The genotype of the GWAS lead variant, rs4921913, is associated with NAT2 protein levels (top) and TG (middle), but shows no association with RNA levels (bottom). (F) Genotype of rs4921913 tags common *NAT2* haplotypes, which encode protein variants linked to varying acetylator phenotypes. All but one *NAT2*4* haplotypes are linked to the C allele at the pSNP, rs4921913. Boxplots show interquartile range (IQR) and median, with whiskers extending to ±1.5 × IQR.

In a second example, a non-coding variant rs4921913 was found significantly associated with the NAT2 protein (p=2.562×10^−12^) in liver tissue, although no eQTL or sQTL were found in tissues examined in GTEx V8 (Fig. 5D-E). This pSNP overlaps GWAS loci associated with a variety of cardiometabolic traits, including triglyceride (TG) and LDLC^45,46^. *NAT2* encodes N-Acetyltransferase 2, an enzyme catalyzing the biotransformation of various drugs and carcinogens^47^. Human *NAT2* is primarily expressed in liver and intestines^48^, with its genomic sequence highly polymorphic among individuals and between populations^49^. Several relatively common protein-coding variants define haplotypes known as *NAT2* alleles^50^. In GTEx liver samples, the protein-increasing rs4921913-C allele was in near perfect LD with haplotypes termed “rapid acetylator” alleles (most commonly, *NAT2*4),* while the protein-decreasing rs4921913-T allele, tagged multiple “slow acetylator” alleles including *NAT2*5B* and *NAT2*6A* (Fig. 5F). These observed patterns of protein abundance variation across haplotypes were consistent with previous immunoblot measurements of NAT2 protein abundance in recombinant expression assays^51^. Examination of specific peptides used for protein quantification indicates that the pQTL reflects changes in protein abundance and cannot be explained by differential detection of peptide variants (Extended Data Fig. 8, Supplementary Information). It has been hypothesized that *NAT2* coding variants may alter protein structure, aggregation, stability and rate of degradation by proteasome; these post-transcriptional mechanisms also reconcile the absence of eQTL evidence^51^. Various mechanisms, through which *NAT2* affects LDLC and TG, have been suggested and include biosynthesis, transport and metabolism^52–54^. Among the 46 genes that were manually curated as likely genes involved in lipoprotein metabolism, transport and remodeling^55^, and whose protein levels were measured in liver, NAT2 displayed the highest positive correlation with HMGCR (r=0.23), SCD (r=0.24) and FADS2 (r=0.25), all of which are associated with cholesterol or fatty acid biosynthesis.

## Discussion

Proteins play a fundamental role as the functional units in nearly all biological and cellular processes. We employed a mass spectrometry-based platform to characterize the inter-individual variation of over 10,000 proteins across five non-diseased tissues. Our findings revealed pervasive inter-individual variation in protein abundance, which are associated with factors such as the donor’s sex, age, cis-RNA expression, and genetic variants. Notably, sex-biased protein expression exhibits high tissue-specificity, except for proteins located on the X chromosome, which mainly represented genes escaping X-inactivation. In contrast, the impact of aging appeared more synchronized across tissues.

The extent to which protein variation can be captured at the transcriptome level has been the subject of a long-standing debate, although previous studies have largely focused on modeling the relationship between mRNA and protein abundance across genes, tissues or cell types^56–58^. The proteomic data generated in our study, integrated with the RNA-seq data from matching tissue samples, offers an opportunity to examine the relationship between transcriptomic and proteomic variation for each gene across individuals. This gene-level RNA-protein correlation across individuals is essential for comprehending the molecular cascades that contribute to the variation of complex phenotypes, as it captures different aspects of biology than the correlations across tissues or genes. Although protein and RNA levels are positively correlated across individuals for the majority of genes, this correlation is generally modest. In other words, RNA variation captures only a small fraction of the observed inter-individual protein variation, which may be attributed to several factors beyond technical measurement error. First, as illustrated by the example of *MIF*, the discordance may be partially due to differences between transcript-level versus gene-level variation. This highlights the importance of accurately quantifying transcript-level gene expression through advancements in computational and experimental techniques. Second, without perturbing RNA abundance, genetic variants can influence protein abundance through post-transcriptional mechanisms, including translation regulation, as well as mechanisms affecting protein stability or degradation as exemplified by *NAT2*. Third, the coordinated regulation of genes in a pathway, at protein or RNA levels, can decouple RNA-protein correlation of a specific gene; this is exemplified by trans-pQTL loci, where protein levels are correlated. Finally, some secreted proteins, but not RNAs, are transported between cells and tissue types and circulate through the bloodstream, further diminishing the RNA-protein correlation. Collectively, our results suggest that transcriptome quantification does not fully capture variation in protein abundance. Instead, integrating transcriptomic and proteomic variation holds the promise of unveiling system-level coordinated regulation and feedback mechanisms governing the dynamics between RNA and protein abundance.

Large-scale GWAS have revealed thousands of genetic variants associated with a wide variety of complex phenotypes and diseases. Yet deciphering genes and the biological processes underlying these GWAS hits has remained a major challenge. Recent methods have seen success in integrating genomic, epigenomic and transcriptomic evidence. We reason that pQTL represents a molecular layer that is more proximal to complex traits and disease phenotypes, and thus can enhance and complement existing strategies. Across the five tissues examined, we identified 1981 cis-pQTL and 30 trans-pQTL. More than 30% of pSNPs overlap with complex trait loci identified through GWAS. Examination of these loci provide important insights. First, an exonic annotation of a GWAS SNP is one of the strongest indicators for functional consequence. Current approaches for linking GWAS SNPs to functional genes strongly favor assigning exonic SNPs to its encoding gene; this strategy ranks as one of the most precise (e.g. highest specificity) assignments in a recent evaluation of computational SNP-to-gene (S2G) linking methods^36^. Yet our results reveal that exonic variants can affect protein abundance of cis-genes other than the ones they encode. Ignoring such possibilities can lead to erroneous interpretation of GWAS associations, as illustrated in the example of variants in *SORT1*, *DPP7* and *AS3MT*. Thus, protein-level variant effects should be considered along with full genomic context. This will likely become more important as the power of mapping pQTL increases. Second, a number of pQTLs, including those at *ABCA8* and *NAT2*, were found at GWAS loci that do not show eQTL evidence. These pGenes represent novel candidate trait-relevant genes, which, in turn, guide further investigation of mechanisms underlying the trait association. In some cases, the discovery of a pQTL may implicate previously unknown gene functions or pathways. Thus, the drug metabolizing gene, *NAT2*, may have a pleiotropic role in the biosynthesis or homeostasis of TG. Collectively, these case studies highlight the value of protein-level information for functional elucidation of GWAS loci. Despite these encouraging examples, it should be emphasized that, just as other molecular QTLs, co-occurrence of pQTL and trait-association alone do not confirm biological causality. Integrating molecular phenotypes at different levels has the potential to nominate and refine candidate genes for functional characterization.

Recent plasma-based population-scale proteogenomic studies have discovered a myriad of cis- and trans-pQTLs, generating important resources for illuminating genes and pathways underlying complex traits and diseases. Our investigation of five distinct human tissues suggests that, while cis-pQTLs and age-related protein variation show substantial overlap between tissues, trans-pQTLs and sex-specific protein abundance hint at tissue-specific genetic architectures. Moreover, a large fraction of proteins are not readily quantified in a single tissue or in blood. Thus, proteogenomic studies that broaden the diversity of tissues and cell types offer a powerful avenue to deepening our understanding of protein abundance regulation, ultimately shedding light on the biology linking genetic variation and human health.

## Methods

### Biospecimen processing and proteomic measurement

The Gene-Tissue-Expression (GTEx) project has built a collection of biospecimens from 49 tissues from 838 post-mortem donors. The biospecimen collection procedure was described in detail in ^59^. The current study focuses on protein expression in five GTEx tissues: sigmoid colon, left ventricle heart, liver, lung, and thyroid. For each tissue, PAXgene-preserved specimens from 144 donors were provided by GTEx Laboratory Data Analysis and Coordination Center (LDACC) at the Broad Institute. The tissues represent donors of both sexes, with ages ranging between 20-70 years (Table S1).

Proteomic data were generated using a tandem mass tag (TMT) 10-plex mass-spectrometry (MS) platform. Each MS run included eight tissue samples from a same tissue and two reference samples; each sample was labeled with a unique TMT-tag. The two references consisted of multi-sample peptide mixtures: one reference, the mega-reference, was derived from 32 organ sites of 14 donors^2^ while the other, tissue-specific reference, represented a peptide pool from all 144 samples from the same tissue as the other eight samples in the same MS run. Peptides were fractionated using liquid chromatography followed by MS using two Thermo-Fisher Orbitrap Fusion Lumos instruments. Mass spectra were searched against the protein-coding transcript translation sequences of GENCODE Release 26 (GRCh38). Protein quantification was based on the ratio of the reporter ion intensity for each sample relative to the tissue-specific reference. Each GTEx tissue specimen was analyzed in two separate runs, with the two resulting protein quantifications later combined. Details of the proteomic experiments, data processing and quality control are provided in Supplementary Information.

### WGS and RNA-seq data

WGS, gene- and transcript-level RNA data are available through dbGaP (Accession ID: phs000424.v8.p2) and GTEx Portal (https://www.gtexportal.org/home/). WGS generated in GTEx (V8) was used for pQTL mapping. For transcriptome-based analysis, normalized and inverse quantile transformed gene-level RNA expression data were used unless otherwise noted. RNA and protein information were paired based on Ensembl gene ID (ENSG). PEER factors based on RNA expression were pre-computed by GTEx; we recomputed genotype PCs on the subset of donors within each tissue. Unless otherwise specified, analyses involving eQTL and sQTL used summary-level statistics produced by GTEx V8^1^.

### Overlap between pQTL and GWAS loci

To investigate the potential downstream effects of pQTL loci in complex traits, we first queried pSNPs identified across five tissues in GWAS catalog^60^ (downloaded on 2024-01-18) and Phenoscanner^61^ (downloaded on 2024-01-18). SNP-trait associations with *p* ≤ 5 × 10^−8^ were retained for further analysis.

### Statistical Analysis

#### Age- and sex-associated proteins

The association between each protein with sex and age was jointly tested using a multivariate linear regression model, adjusting for ischemic time, genetic and protein PCs as covariates. All proteins that were quantified in at least 72 donors were tested. We chose, *a priori*, to include the top three genetic PCs and top protein PCs that explained at least 5% of the total protein phenotypic variance in a tissue. This leads to the inclusion of three, four, four, four and three PCs for colon, heart, liver, lung and thyroid, respectively. The q-values were calculated within each tissue; an FDR threshold of 0.1 was used to define age- or sex-associated proteins. To make the effect sizes comparable for visualization in Fig. 2D-E, sex and age variables of the donors were standardized within each tissue to have a standard deviation of one. The sharing of sex- and age-association with protein abundance across tissues was evaluated using an empirical Bayes approach, implemented in the R package MASHR (Supplementary Information).

#### Protein-RNA and protein-protein correlations

Pearson correlations between RNA and protein abundance of each gene, as well as between the protein abundance of a pair of genes, were calculated in each tissue separately, requiring proteins quantified in at least 72 specimens. Covariate adjustments for RNA and proteins, as well as statistical methods for testing protein-protein correlations within and across complexes are described in Supplementary Information.

#### cis-pQTL and trans-pQTL mapping

Both cis- and trans-pQTL mapping was performed in each tissue separately. Autosomal encoded proteins that were quantified in at least 72 donors with WGS in a tissue were included in pQTL mapping. We used a linear model to assess the association between genotype and quantile-transformed protein phenotypes, adjusting for the following covariates: sex, age, ischemic time, the first 3 genotype PCs and the same number of top protein PCs as described in the sex- and age-association analyses. Genetic variants from GTEx V8 were filtered to retain variants with minor allele frequency (MAF) ≥ 0.05 among individuals with proteomic data; the genotype PCs were recalculated using genotype data of the 144 donors in each tissue.

For cis-pQTL mapping, variants within a window of 1Mb upstream and downstream of the protein coding region were tested. The significance testing procedure for cis-pQTLs is analogous to that used for GTEx cis-eQTL mapping. Briefly, for each protein phenotype, a null distribution of the minimum p-value across all SNPs was derived by permuting the protein phenotypes relative to genetic variants, so that the correlation between variants (linkage disequilibrium, LD) were preserved. Subsequently, proteome-wide false discovery rate was calculated using gene-level permutation p-values. An FDR < 0.1 was defined as significant cis-pQTL (Table S5).

We defined trans-region as variants located more than 1Mb up- and down-stream of the protein-coding sequence or on a different chromosome. We used the same model, covariates, as well as protein and variants inclusion criteria as for cis-eQTL analysis (described above). Following the practice of the GTEx trans-eQTL mapping, a gene-level adjusted p-value was calculated by taking the smallest nominal p-value per gene across all tested SNPs and multiplying that p-value by 10^6^ and truncated at 1. Lastly, the gene-level adjusted p-values from all protein phenotypes were pooled to compute the tissue-wide FDR via R package, q-value.

#### Replication of cis-pQTL

We used two external data sets to validate the cis-pQTLs reported here: (1) A plasma-based study from INTERVAL study, which used an aptamer-based multiplex protein assay (SOMAscan) to quantify 3,622 plasma proteins in 3301 healthy blood donors^8^, and (2) the Fenland study, which used a combination of SOMAscan and Olink arrays to quantify a total of 4,775 proteins in 12,084 participants^9^. We took unique pairs of SNP-gene discovered in each GTEx tissue and queried their corresponding association p-values in INTERVAL and Fenland summary statistics, respectively. The replication was quantified using the π_1_ statistics, which estimated the non-null proportions of GTEx pQTL in each of the replication datasets (Supplementary Information & Extended Data Fig. 7).

#### Mediation analysis for genetic effect on protein abundance via mRNA

We performed a mediation analysis to formally test the hypothesis that the pSNP-pGene association was mediated through regulation at the RNA expression level. At each pQTL, the mediation analysis compared two linear models with the protein abundance as the outcome. Both models included the genotype of pSNP; one model further included the pSNP-predicted RNA abundance while the other did not. The mediation effect was assessed by the difference in the coefficients corresponding to the pSNP genotype between the two models (Supplementary Information & Fig. S12).

## Data Availability

Normalized protein abundance and pQTL call sets will be distributed through the GTEx Portal: https://gtexportal.org. Raw proteomic data will be deposited to ProteomeXchange Consortium via the PRIDE partner repository. GTEx individual-level WGS and RNA-seq data are available through dbGaP (Accession phs000424.v8.p2).

## Code Availability

Scripts for protein normalization and pQTL mapping will be available at https://github.com/tanglab/

## Acknowledgements

We thank the donors and their families for their generous gifts of organ donation for transplantation, and tissue donations for the GTEx research project, and J. Struewing for his support and leadership of the GTEx project. We thank Fengchao Yu and Hui-Yin Chang for assistance with protein quantification analyses and data transfer. Sophie Candille and Peng Ding for helpful discussions. Kristin G. Ardlie, François Aguet, Ellen Gelfand and the GTEx LDACC are gratefully acknowledged for providing GTEx tissue samples. We thank investigators of Fenland and INTERVAL for providing full summary-level pQTL mapping statistics, which enabled replication analyses.

## Author contributions

H.T. and M.P.S. conceived the study; H.F led all computational and statistical analyses supervised by H.T.; L.J. led the TMT-MS proteomics experiments with the contribution from R.J. and J.C.; F.V. and A.I.N performed peptide and protein identification and quantification analyses and advised on peptide spectra analyses; D.G. and T.L. contributed long-reads data. T.L., A.P.R, M.P.S. and L.J. contributed to data analyses and interpretation; H.F. and H.T. lead the writing of the manuscript and supplements, with detailed comments from T.L., A.P.R., A.I.N. and M.P.S.. All authors read and approved the final manuscript.

---

**Extended Data Figure 1.**
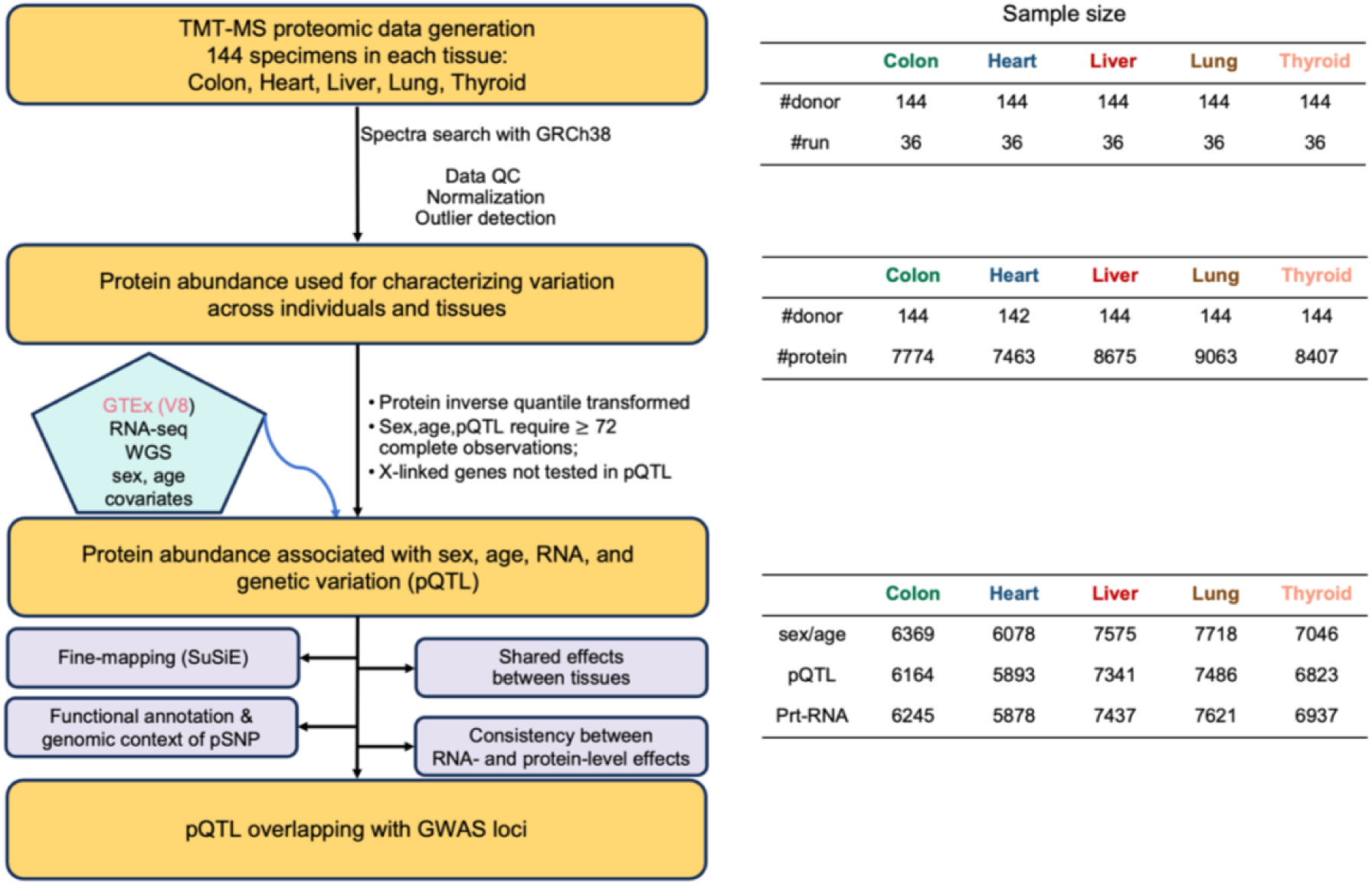
Overview of study design and a summary of sample size / number of proteins used in each analysis.

**Extended Data Figure 2.**
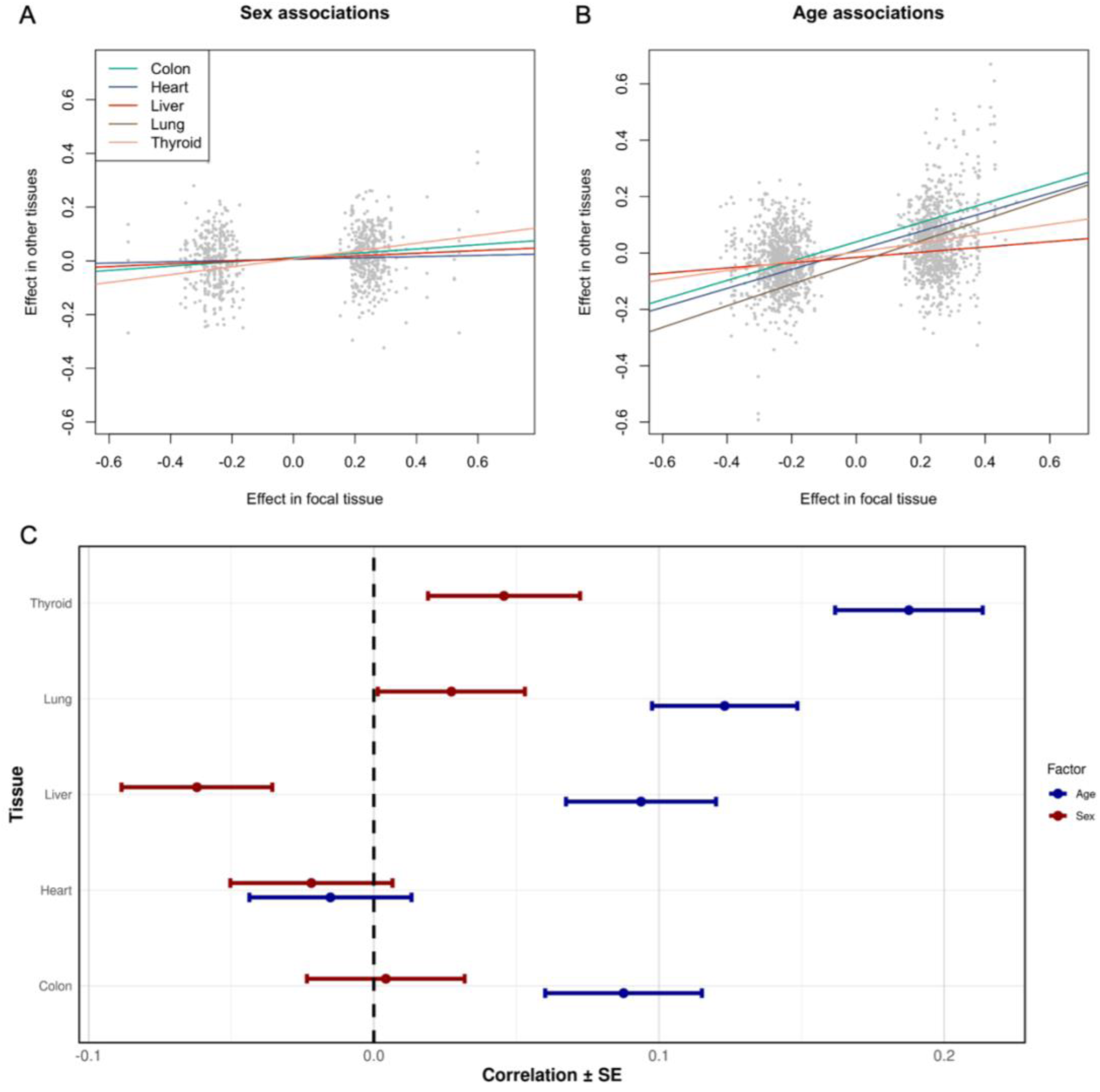
Heterogeneity of effects of sex and age on protein abundance across tissues. (A-B): Treating one GTEx tissue as a focal tissue, the effect sizes of significant sex-associated (A) or age-associated (B) proteins (FDR<0.1) in the focal tissue are plotted along the x-axis; the corresponding effect sizes in other tissues are plotted along the y-axis, regardless of the statistical significance. The focal tissue is liver in (A) and Thyroid in (B). Generalizability of the effects of sex (or age) across tissues is quantified with a linear model that uses the effect sizes in each focal tissue to predict the corresponding effects in non-focal tissues, adjusting the non-focal tissues as a random effect. The colors of the regression lines indicate the focal tissues. For the sex-associated proteins, X-linked proteins are excluded from the regression. (C) The estimated effects of sex (red) and age (blue) were compared between a GTEx tissue and plasma. Points indicate Spearman correlation, with the error bar indicating standard error. The effects of sex and age on plasma proteins were taken from the UK Biobank Pharma Proteomics Project (UKB-PPP)^7–9^ (Supplementary Table (ST5)). The correlation was computed based on proteins quantified in both UKB-PPP and the GTEx tissue under consideration; this resulted in 1388, 1314, 1509, 1590 and 1487 proteins for the comparisons with GTEx colon, heart, liver, lung and thyroid, respectively.

**Extended Data Figure 3.**
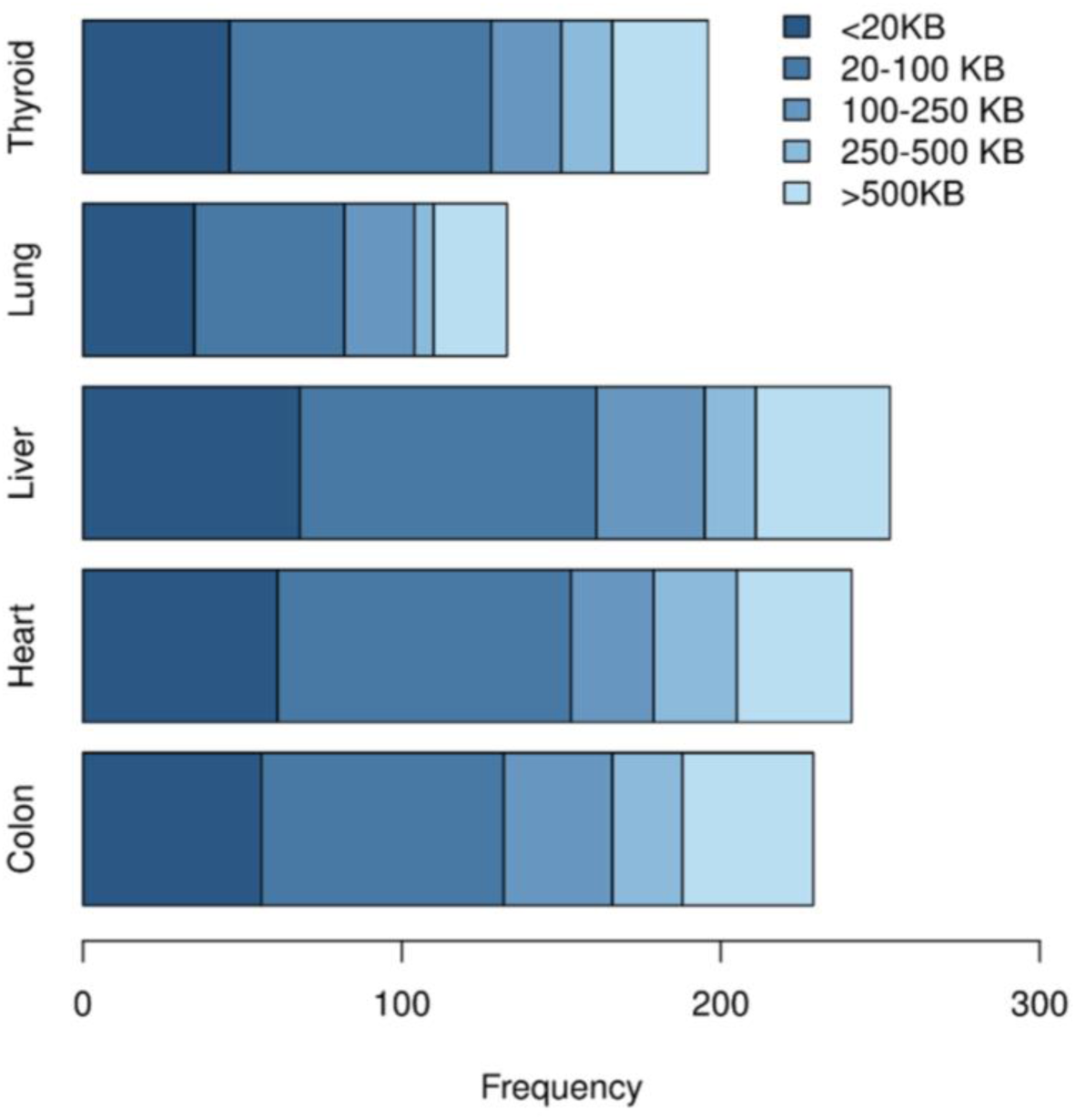
Distribution of physical distances (in bp) between the pSNP and TSS of the pGene at each pQTL, whose pSNP was located outside of the pGene.

**Extended Data Figure 4.**
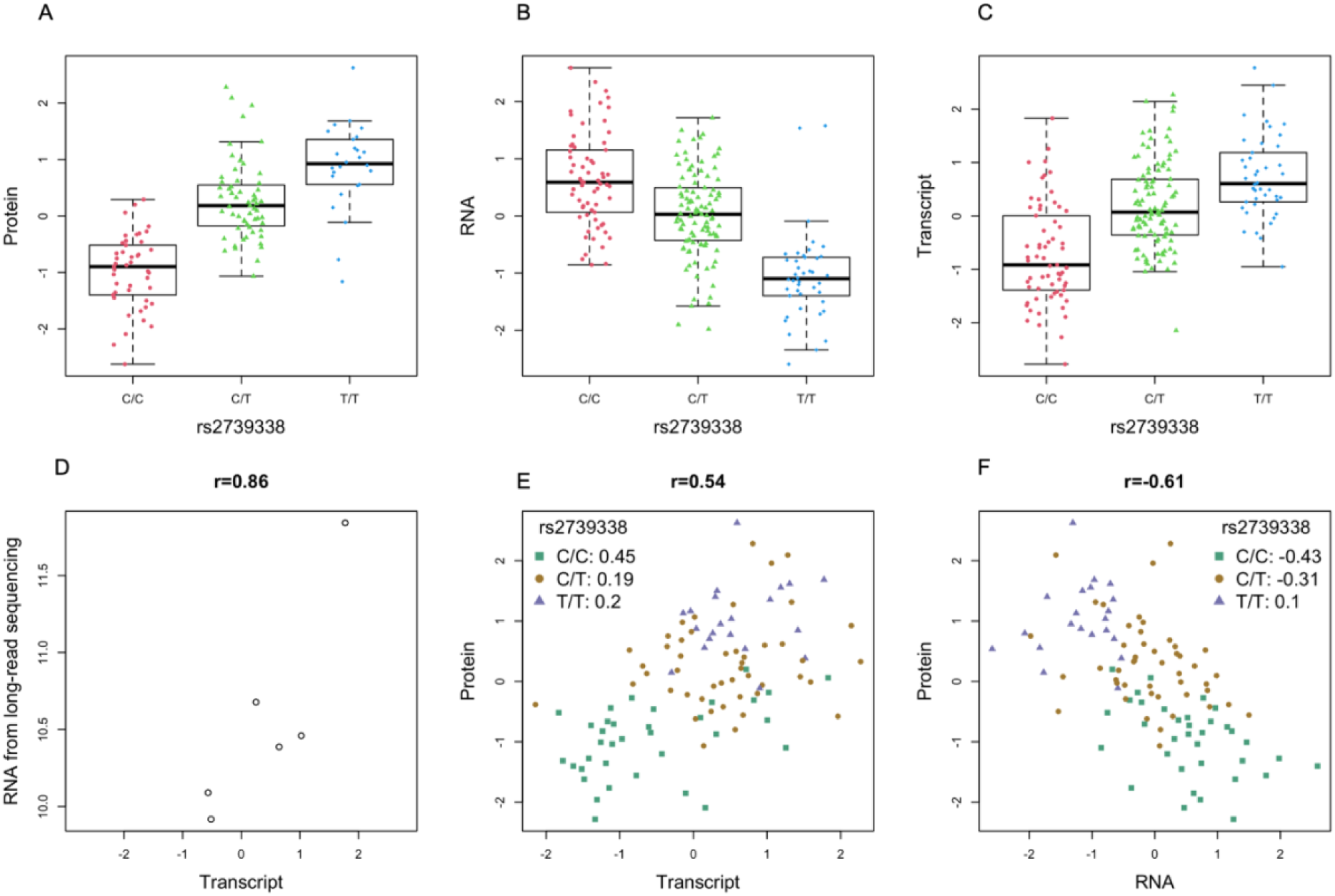
Transcript QTL (trQTL), but not gene-level expression QTL (eQTL), supported pQTL of MIF in liver. (A) rs2739338-T was associated with increased MIF protein (*p* < 9.2 × 10^−18^). (B) rs2739338 was associated with gene-level expression, but with an opposite effect (*p* = 3.6 × 10^−29^). (C) rs2739338-T was associated with an increased transcript level of ENST00000215754, which encodes the protein (*p* = 5.18 × 10^−32^). (D) Transcript level of ENST00000215754 in six liver samples, which were quantified both with long-read sequencing (y-axis) and with RNA-seq in GTEx V8 (x-axis). (E) MIF protein level (y-axis) is positively correlated with ENST00000215754 transcript level estimated based on GTEx V8 RNA-seq (x-axis). (F) MIF protein level (y-axis) is negatively correlated with gene-level RNA abundance (x-axis). More surprisingly, each of the three MIF transcripts is inversely correlated with the gene-level RNA abundance. Boxplots in (A-C) show interquartile range (IQR) and median, with whiskers extending to ±1.5 × IQR.

**Extended Data Figure 5.**
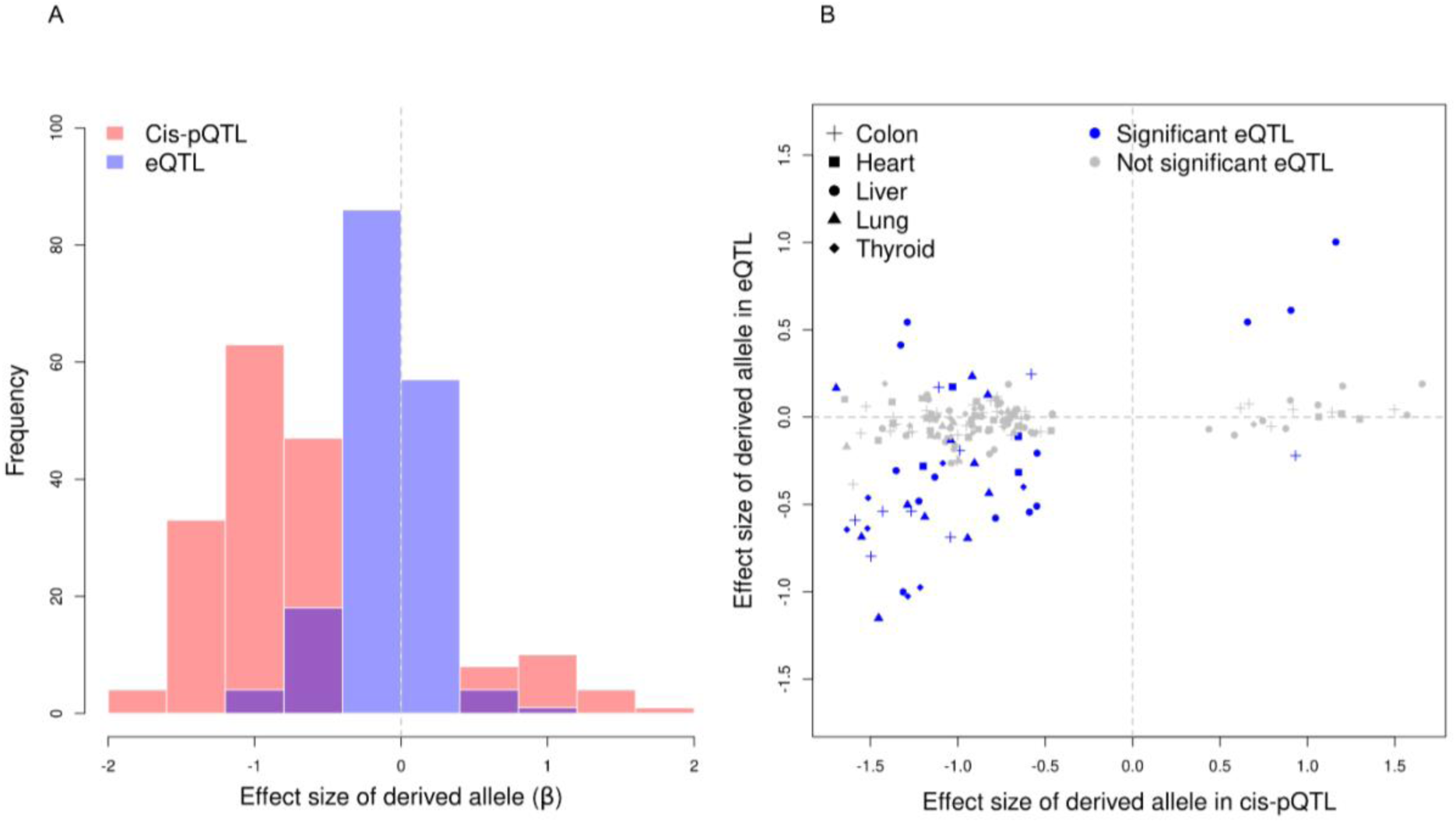
Allelic effects at 172 pQTLs, where the pSNP represented protein altering variants (PAV). (A) Histogram of the non-reference allelic effects of PAV on protein (pink) and RNA (blue) abundance. The non-reference alleles were associated with a decreased protein level at 87% sites and decreased RNA level at 64% sites. (B) Effect size and direction of the non-reference alleles on protein (x-axis) and RNA (y-axis). A pSNP that is a significant eQTL of the same gene is indicated in blue; otherwise, it is colored in grey.

**Extended Data Figure 6.**
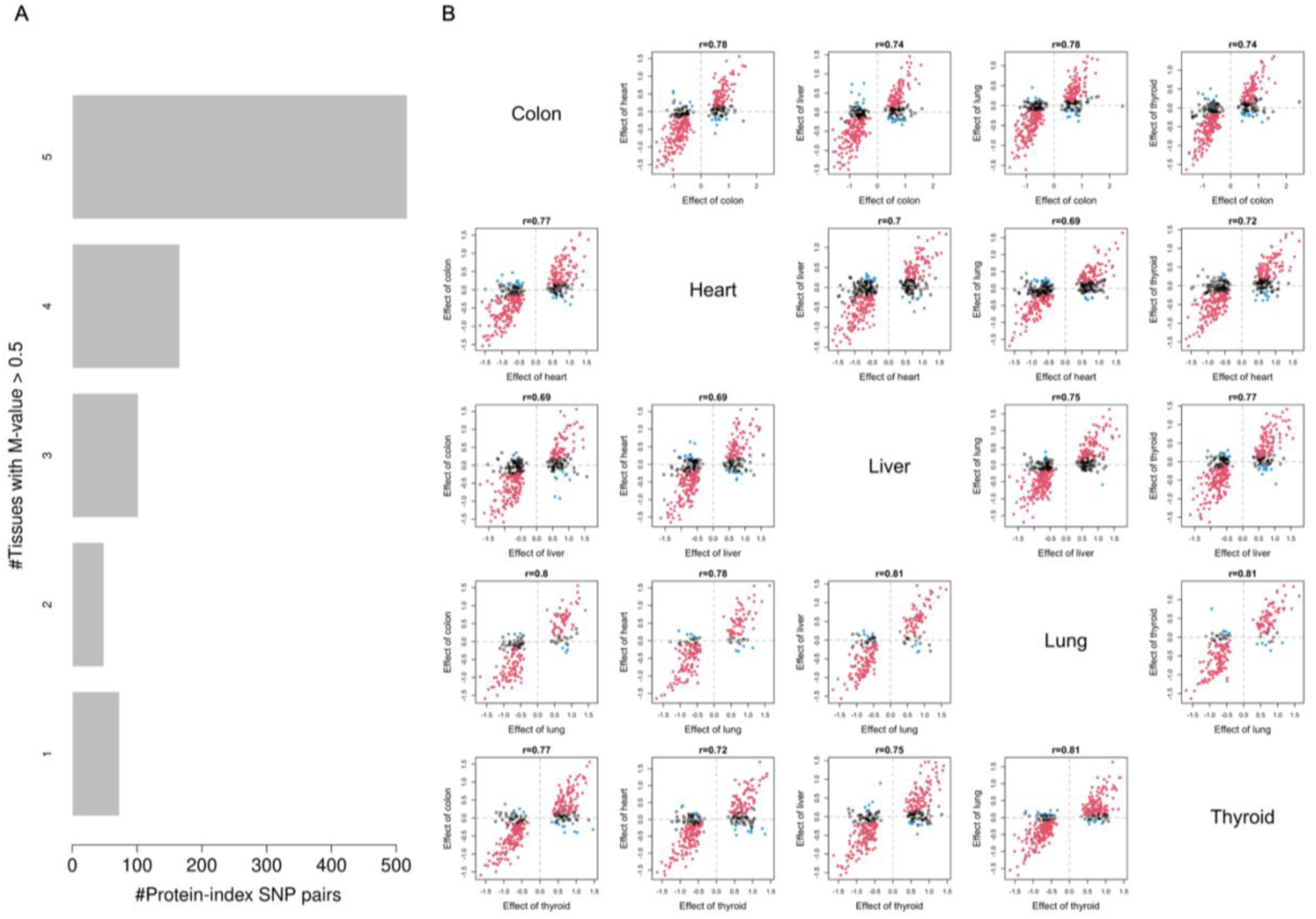
Tissue-sharing of pQTLs. (A) Number of tissues in which a pQTL was active, estimated using a Bayesian meta-analysis approach (Supplementary Information). (B) Joint distribution of allelic effects in pairs of tissues. pSNPs of all pQTLs detected in one tissue (x-axis) were included. Their corresponding allelic effect size in another tissue (y-axis) were categorized as consistent (same direction and significant, red), inconsistent (opposite direction and significant, blue), or not significant (black). The number in the title of each subplot represents Spearman correlation between the estimated allelic effect in the two tissues.

**Extended Data Figure 7.**
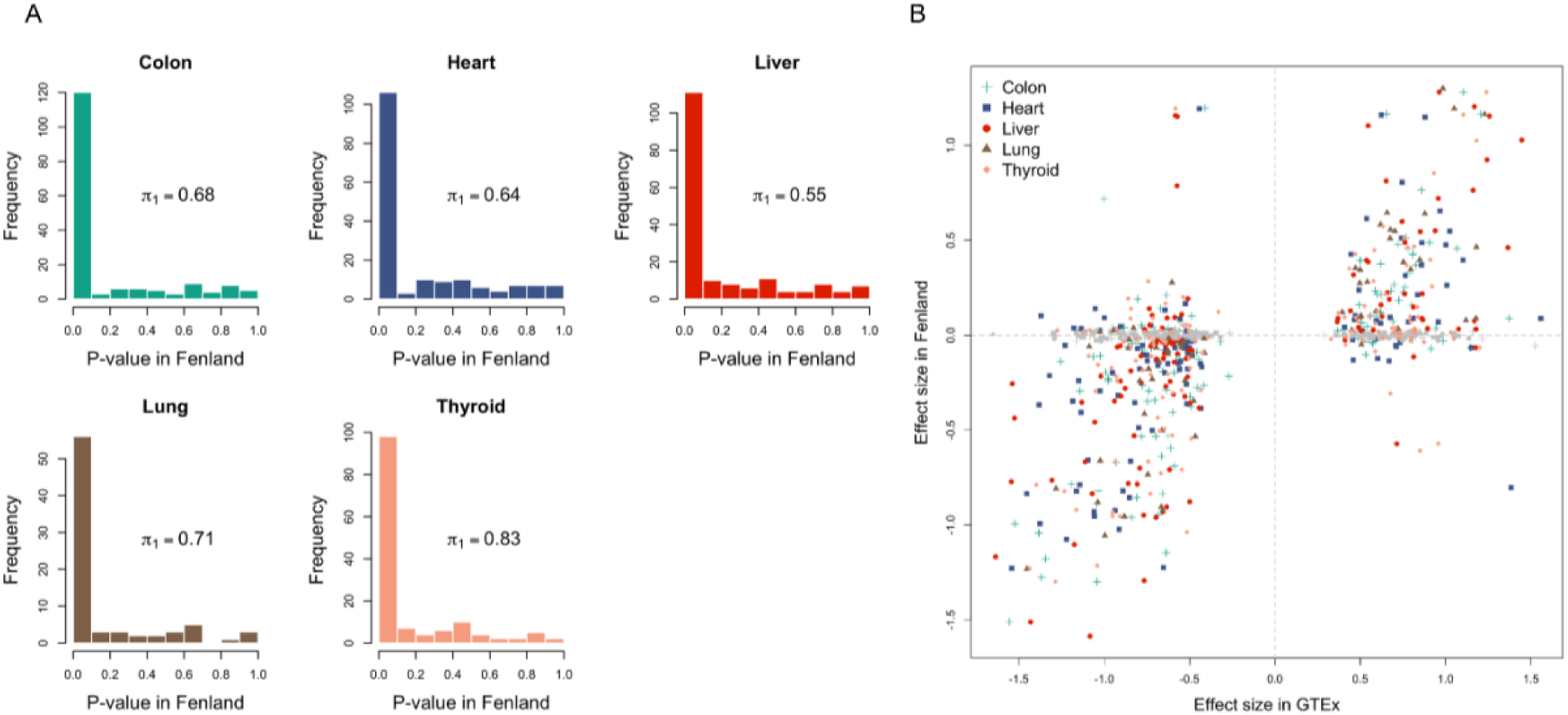
Replication of GTEx tissue cis-pQTLs in Fenland Plasma Study. For pQTLs discovered in each GTEx tissue, the corresponding SNP-protein association statistics were queried in Fenland summary statistics. (A) Distribution of p-values in Fenland; *π*_1_ is a measure of replication rate. (B) Joint distribution of the estimated effect size of tissue pQTL (x-axis) and the corresponding allelic effects in Fenland. Each point represents a pQTL discovered in GTEx; symbol and color of the point indicate the tissue of discovery, unless the pQTL is not replicated in Fenland (FDR>0.1), in which case the point is colored as gray.

**Extended Data Figure 8.**
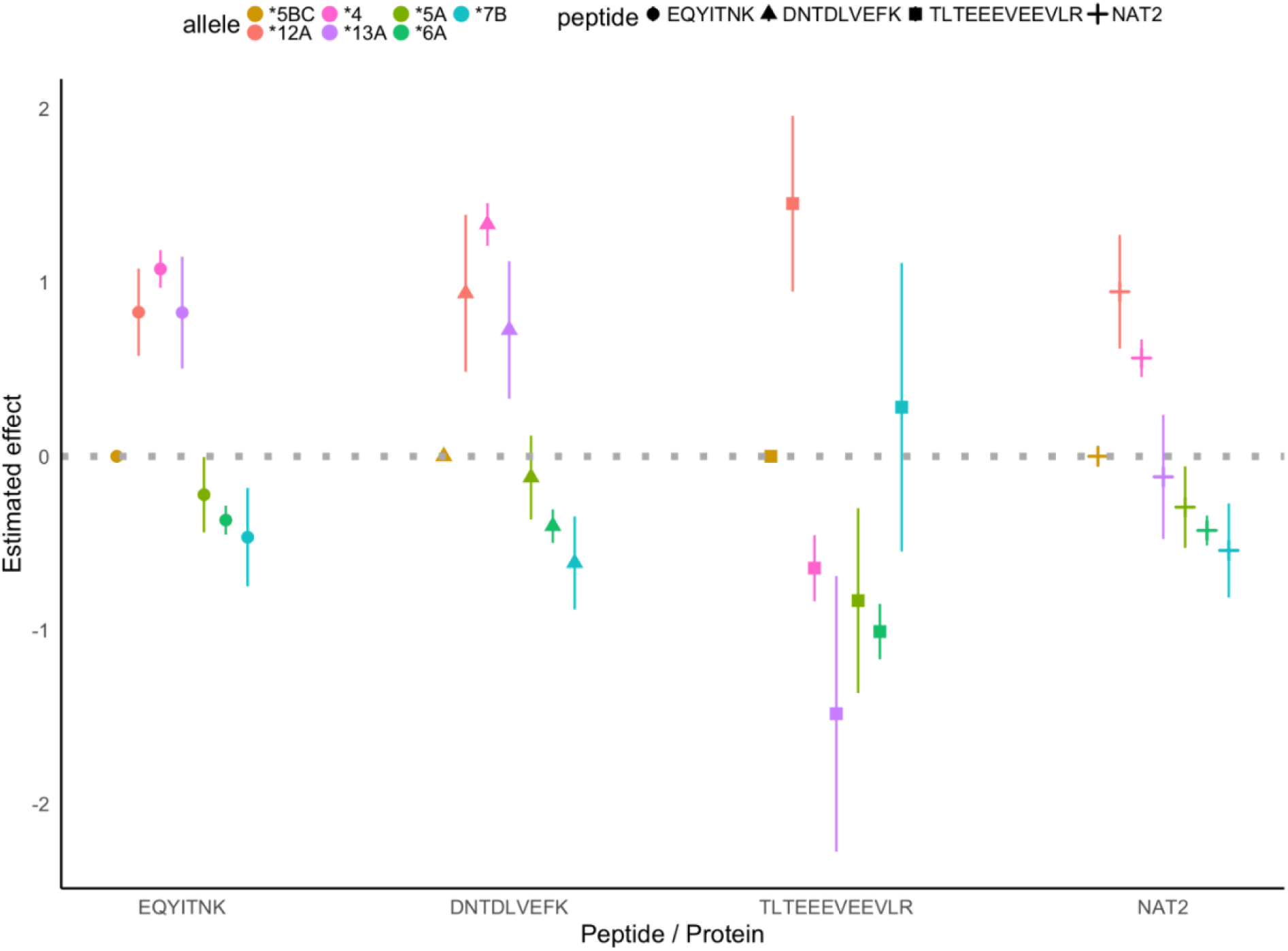
Estimated *NAT2* haplotype effect on the abundance of three NAT2 peptides, which were quantified by mass-spectrometry and used for NAT2 protein quantification (Supplementary Information). The group labelled “NAT2” denotes protein quantification. Peptides EQYITNK and DNTDLVEFK do not overlap with any common coding variants (MAF>5%) in our sample. Peptide TLTEEEVEEVLR harbors a coding variant, rs1208. The reference peptide sequence used for spectra search is the same as haplotypes *NAT2*5B*, *\*5C* and *\*12*. *\*5B* (N=82) and *\*5C* (N=1) were combined and used as the reference group in the haplotype effect estimation. The protein-increasing pSNP allele, rs4921913-C, is in high LD with *NAT2*4C*. Center and the segments represent point estimates and ±1 standard error of the estimated haplotype effects, which were the coefficient estimates using a linear regression model, with peptide / protein abundance as the response, treating haplotype as a categorical variable. The sample sizes for peptides EQYITNK, DNTDLVEFK, TLTEEEVEEVLR and gene NAT2 were 120, 200, 158 and 249, respectively.

