## Supplementary Information for "Regulation of protein abundance in normal human tissues"

### Supplemental Information

#### Table of Content

**1. TMT-MS proteomics experiments**
**2. Proteomic data processing and quality control**
**2.1 Protein identification and quantification**
**2.2 Normalization and combining replicates data**
**2.3 Quality control assessment and outlier removal**
**2.4 Missing rate and its impact on analyses**
**3. Factors associated with protein abundance**
**3.1 Sharing of protein abundance with sex and age across tissues**
**3.2 Gene set and complex definition**
**3.3 Correlation between protein and mRNA abundance**
**3.4 Protein-protein correlations in complexes**
**4. Protein QTL (pQTL) mapping**
**4.1 Phenotype and covariates used in cis-pQTL and trans-pQTL mapping**
**4.2 cis-pQTL mapping**
**4.3 trans-pQTL mapping**
**4.4 Fine-mapping at cis-pQTL with SuSiE**
**4.5 Replication of cis-pQTL**
**4.6 Tissue-sharing of cis-pQTL**
**5. Genomic and transcriptomic context of cis-pQTLs**
**5.1 Functional annotation of cis-pQTL**
**5.2 Gene-specific functional annotation**
**5.3 eQTL-mediated pQTL**
**5.4 Defining eQTL-supported cis-pQTLs**
**5.5 Transcript-level QTL (trQTL) mapping**
**5.6 Additional information of MIF example**
**6. Additional analyses of NAT2**

**7. Gene ontology (GO) enrichment analysis**

#### 1. TMT-MS proteomics experiments

This study analyzed protein expression biospecimens derived from five GTEx organ sites: sigmoid colon, left ventricle heart, liver, lung, and thyroid. Henceforth, we refer them as tissues with abbreviated names colon, heart, liver, lung and thyroid. The biospecimen collection procedure was described in detail in [1]. For each tissue, PAXgene-preserved specimens from 144 donors were provided by GTEx Laboratory Data Analysis and Coordination Center (LDACC) at the Broad Institute. The tissues represent donors of both sexes, with ages ranging between 20-70 years (Table S1).

Proteomic data were generated using a tandem mass tag (TMT) 10-plex/MS3 strategy, which analyzed 10 isotopically labeled samples in a single mass-spectrometry (MS) experiment[2]. The TMT-labeled 10-plex mixture was extensively fractionated through a two-dimensional liquid chromatography (LC) to increase the proteome coverage. Sample preparation, protein extraction and digestion, TMT-labelling, LC separation and MS data acquisition followed methods described in [3] with the following modification (illustrated in Fig. 1A). Because the primary focus of this study was to characterize protein variation across individuals within a tissue, samples from a given tissue were analyzed separately from other tissues. Two pooled reference samples were used. The mega-reference, labelled with TMT-126, was the common reference samples used in Jiang et al. and represented a peptide mixture from 32 organ sites derived from 14 donors[3]. In contrast, the tissue-reference (labelled with TMT-131), was formed by pooling an equal amount of protein extracted from all 144 specimens of a specific tissue. Each MS run (abbreviated as "run" subsequently) included the mega-reference, a tissue-reference, and eight individual tissue samples from the same organ site as the tissue-reference. The grouping of the eight samples in a run was randomized. Each specimen was analyzed in technical replicates, which had the same TMT-tag and were analyzed in two different runs. In total, 36 TMT 10-plex runs were performed in each tissue, of which 288 samples corresponded to single-donor specimen and the remainder represented references. As described below, protein ratios were computed relative to the tissue-specific reference; the inclusion of the mega-reference sample enabled data quality check and improved normalization across mass-spectrometry runs.

#### 2. Proteomic data processing and quality control

##### 2.1 Protein identification and quantification

Peptide and protein identification and quantification were performed using MSFragger, followed by the Philosopher pipeline (https://philosopher.nesvilab.org/) and TMT-Integrator (http://tmt-integrator.nesvilab.org/)[4–6]. MS/MS spectra were searched against the protein-coding transcript translation sequence of GENCODE Release 26 (GRCh38), appended with an equal number of decoy sequences and common contaminants (www.gencodegenes.org/human/release_26.html). Peptide to spectrum matches (PSM) from all 36 TMT 10-plex experiments for each tissue were processed together to assemble peptide into proteins. The quantification of protein was based on the intensities of TMT reporter ions from the MS3 scan. Specifically, in each TMT 10-plex run, the intensity of each PSM in each TMT channel was log2 transformed, and the reference channel intensity was subtracted from that for the other nine channels, thus generating log2-based "PSM-ratios" (relative to the reference). The gene-level relative protein abundance is defined as the median PSM-ratio among all PSMs (after removing outliers) mapped to the gene. A reference intensity of the protein was produced based on the MS1 intensity of the corresponding precursor-ions. Additional details of protein identification, quantification, filtering, and the parameters used in these steps followed the whole proteome analysis of [7]. Over 60% of the proteins quantified in the current study were not quantified in the recent, plasma-based, proteogenomic study of the UK Biobank[8] or Fenland[9] (Fig S1).

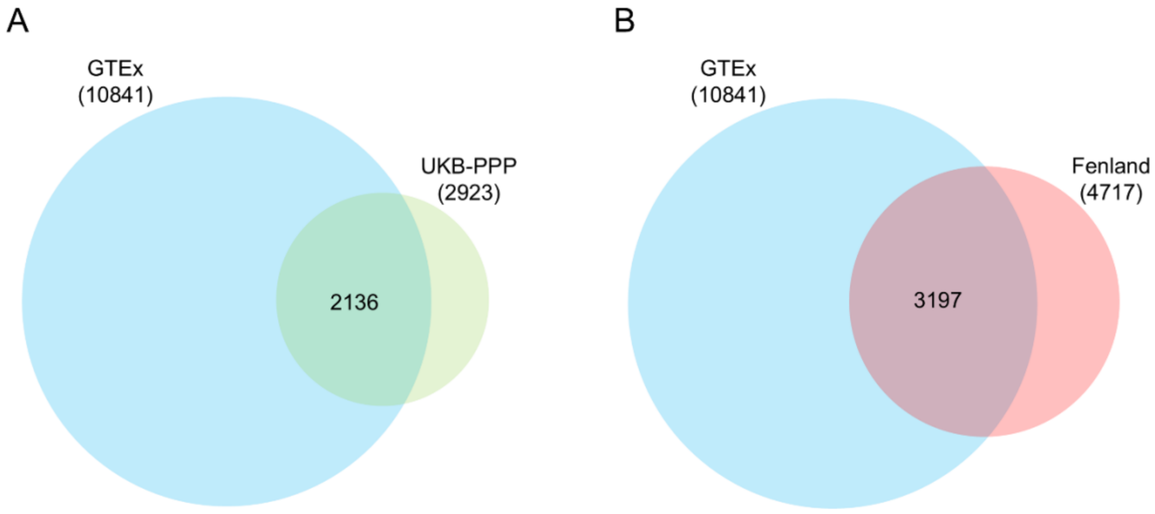


Fig. S1. Comparison of proteome coverage. The number is for the total quantified proteins. Shared and unique proteins quantified in current study, compared to those measured in the UK Biobank Plasma proteomic study (A) and those in the Fenland study (B). Area of the circles are proportional to the number of proteins.

#### 2.2 Normalization and combining replicates data

Let $Y_{gk}$ be the $\log_2$ protein abundance ratio (which we refer to as "protein abundance") of gene $g$ in a sample $k$ relative to the reference sample of the corresponding run. This is the output from the quantification procedure described in the proceeding section, and this quantity can be modelled as a measurement with stochastic errors that arose from factors related to tissue sample, the run, and the protein. We developed a normalization method, ProMix[10], which implemented a linear mixed model that accounted for these factors:

$$Y_{gk} = \mu_g + \alpha_k + b_{gr_k} + \gamma_{gi_k} + e_{gk}.$$

The term, $\mu_g$ denoted the overall deviation across all samples from the reference for protein $g$. The term, $\alpha_k$, described the deviation of sample $k$ from the run-specific reference across all proteins, and accounted for the variation in total proteins in a sample. Let $r_k$ denote the mass-spectrometry run that included sample, $k$. The random-effect term, $b_{gr_k} \sim \mathrm{N}(0, \sigma_{gr}^2)$, represents the run-level variation of protein $g$. Intuitively, this can be interpreted as the measurement error of the protein in the reference in run, $r_k$. The term, $\gamma_{gi_k}$, is the primary variable of interests and represents the specimen-effect of protein, $g$, in specimen, $i_k$. The estimate, $\hat{\gamma}_{gi_k}$, is used as the normalized protein abundance in subsequent analyses. Finally, $e_{gk} \sim \mathrm{N}(0, \sigma_{ge}^2)$ is the residual term for gene $g$ in sample $k$. This method also takes advantage of the technical replicates and a second reference sample (mega-reference), which was included in each run. When a protein is quantified only in a small fraction of samples, the linear mixed model implemented in ProMix may be numerically unstable, yielding noisy estimates of run effects ($b_{gr_k}$). We implemented a number of *post hoc* quality assessment metrics to identify proteins with unreliable estimates; for these proteins the run-effects were shrunken to 0, effectively reverting to a standard normalization approach in mass-spectrometry-based proteomics and metabolomics analyses called probability quotient normalization (PQN) approach, which set the median relative protein abundance across all quantified proteins in a sample to zero[11]. Specifically, PQN was used for when (1) protein abundance was measured in less than 30 samples or less than 10 complete technical replicates; or (2) adjusting the estimated run-effects increased (rather than decreased) the mean squared error (MSE) between replicates; or (3) adjusting the estimated run-effects increased (rather than decreased) variation across specimens.

As a last step, to reduce the influence of skewness and outliers in the protein abundance, an inverse normal transformation was applied for each protein, $g$, in parallel to the eQTL mapping procedure used in GTEx:

$$Z_{gk} = \Phi^{-1}\left(\frac{\mathrm{rank}(\gamma_{gk}) - 3/8}{\mathrm{K} + 1/4}\right),$$

where $\mathrm{rank}(x)$ denoted the rank of $x$ among $K$ observations. The resulting quantities were used as the protein phenotypes for the assessments of sex- and age-association, for pQTL mapping, as well as for computing RNA-protein and protein-protein correlations. Descriptive statistics of all quantified proteins can be found in Table S2.

**2.3 Quality control assessment and outlier removal**

Prior to normalization and combining protein abundance measured in the two replicates of each biospecimen, we performed preliminary analyses to identify abnormal measurements.

For these analyses, protein quantification was normalized using PQN. Hierarchical clustering of all samples within each tissue confirmed that replicates derived from the same donor were almost always clustered together (Fig. 1B and Fig. S2).

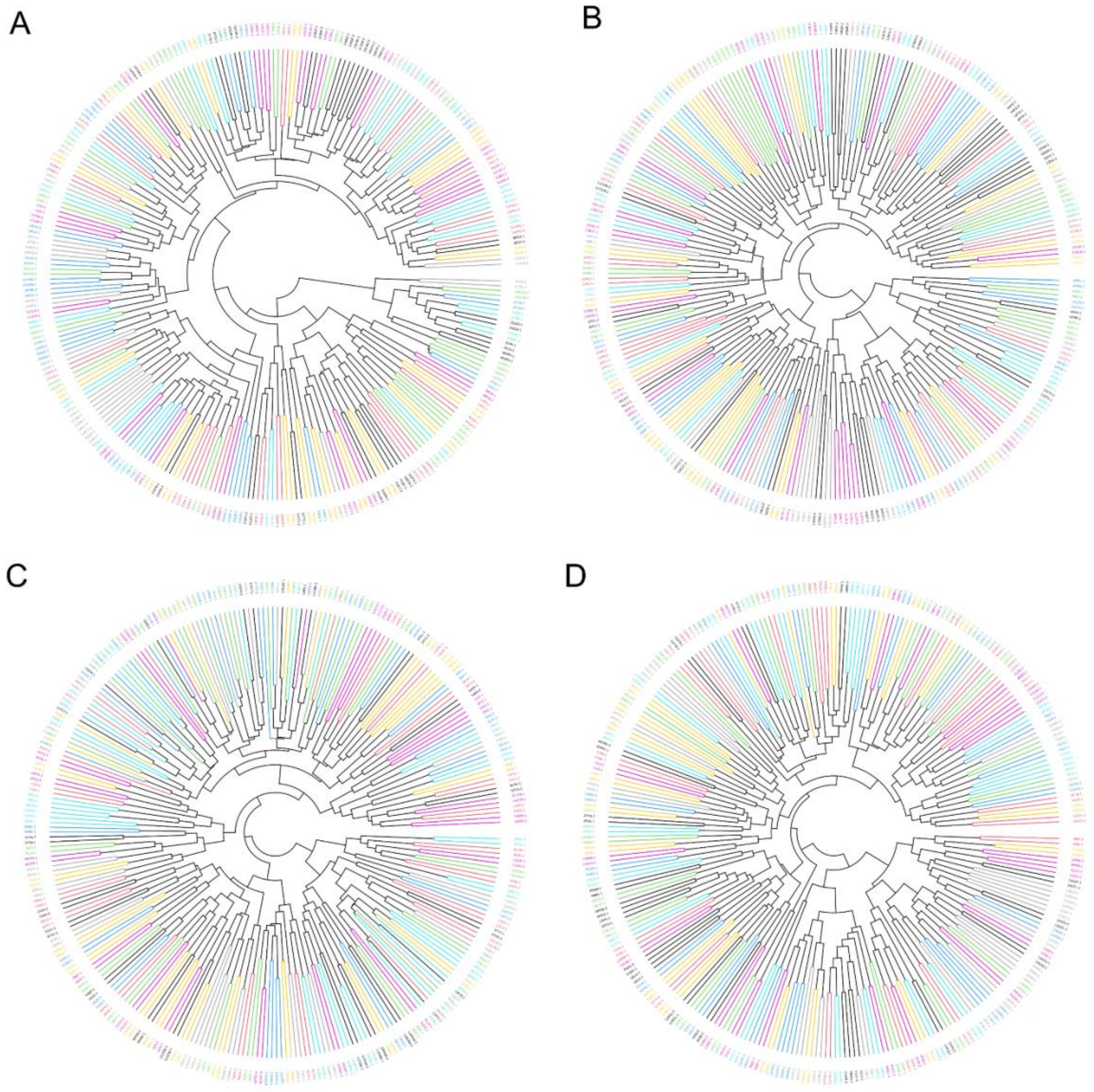


Fig. S2. Hierarchical cluster plot technical replicate samples of (A) colon; (B) heart; (C) lung and (D) thyroid. Plot of liver is shown in Fig. 1B. Replicates of the same tissue specimens (e.g. same donor), which were always analyzed in different runs, are colored the same; 93% of all pairs cluster together.

Next, principal components analysis was performed for each tissue separately to identify anomalous specimens or samples. In colon, the mega-reference in one run appeared as an outlier compared to that from the other 35 runs. In heart, a total of four samples, representing two replicate pairs, were detected as outliers based on PC1 and PC2. These outliers were removed prior to subsequent analyses (Fig. S3). Thus, the final proteomic data represent 144 tissue specimens in each of colon, liver, lung and thyroid, and 142 tissue specimens in heart.

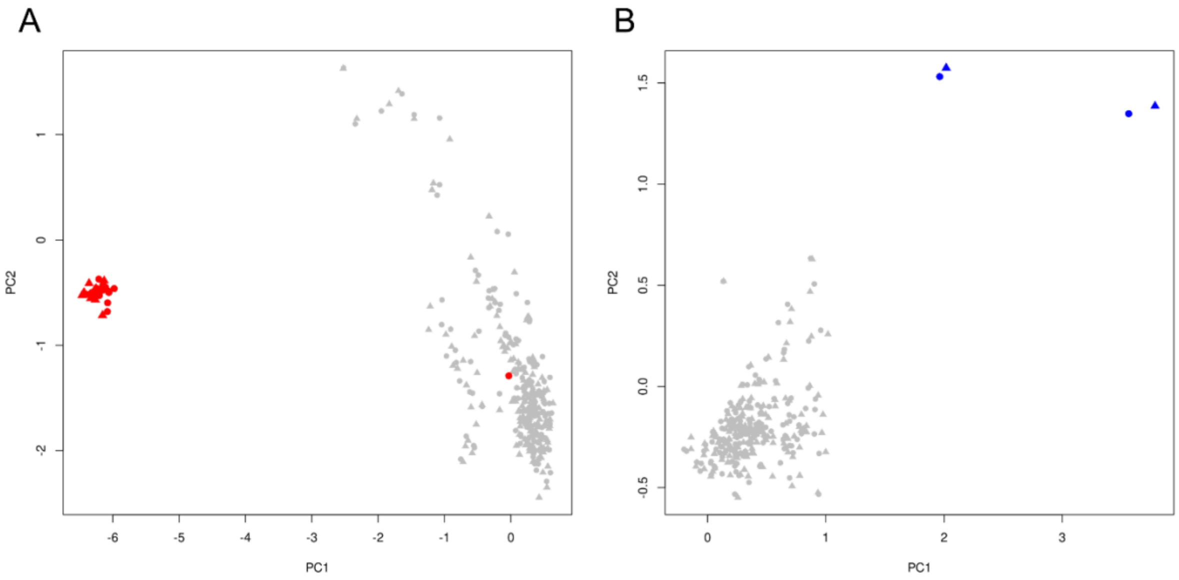


Fig. S3. PCA at sample level detecting (A) an outlier in colon (representing a mega-reference in one run; red points are mega-references from the 36 runs) and (B) four outliers in heart (mega references removed in PCA), corresponding to the replicate pairs of two specimens. These outliers were excluded from subsequent analyses.

#### 2.4. Missing rate and its impact on analyses

The missing rate of a protein depends on protein abundance as well as the ionization properties of peptide fragments. It is well-recognized the large dynamic range spanned by the proteome poses a challenge for untargeted protein quantification. Low-abundant proteins may incur higher missing rate or be less accurately quantified[12]. In our data, most missing occurs at a run level. Since biospecimens were randomly assigned to MS runs with respect to donor characteristics, missing observations generally do not inflate type I errors (false positive discoveries) in single-protein analyses, such as the identification of proteins associated with sex, age or genetic variants. However, statistical power is reduced in analyses of proteins with higher missing rates or higher measurement errors. Thus, we restricted association testing to proteins that are quantified in at least 72 (≥ 50%) donors in a tissue. Fig. S4 summarizes the missing rates in five tissues.

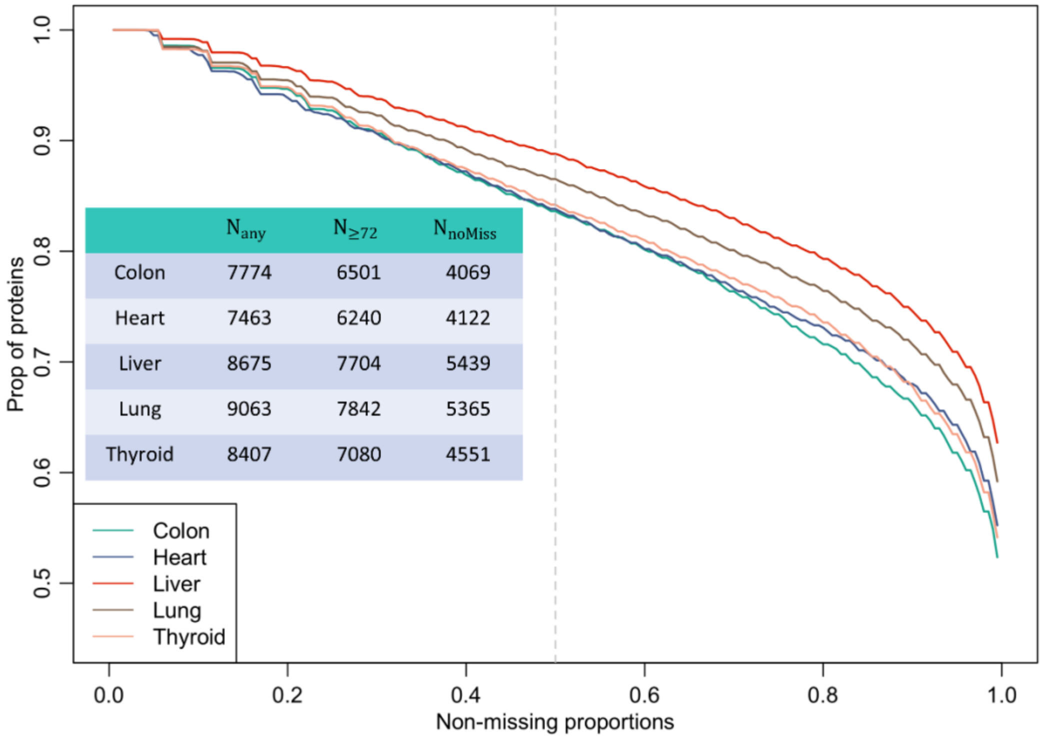


Fig. S4. Proportions of proteins (y-axis) exceeding specific non-missing threshold (x-axis). Inset table summarizes the total number of proteins ($N_{any}$), number of proteins with more than 72 non-missing observations ($N_{\geq 72}$) and the number of proteins quantified in all specimens ($N_{noMiss}$). Note that the actual numbers of proteins used for age- and sex-association analyses are 6369, 6078, 7575, 7718 and 7046 for colon, heart, liver, lung and thyroid, respectively. The corresponding numbers of proteins used for pQTL testing are 6164, 5893, 7341, 7486 and 6823 (Extended Data Fig. 1). The difference is due to missing covariates and genotype data; for pQTL, proteins on the X chromosomes were further removed.

For analyses across proteins, such as the comparison of inter-donor variation between tissue-shared and tissue-specific proteins, missing rate and measurement error can act as confounding factors. As the reference ion intensity of a protein is influenced by protein abundance, detection rate and measurement error, it serves as a surrogate variable for the confounding factors. The overall reference intensity of a protein was estimated by the mean of reference intensity in each run, which was, in turn, computed as a weighted sum of the

MS1 intensities of the top 3 most intense peptide ions quantified for that run. The reference intensity value was not used in computing protein abundance ratio; however, this quantity was correlated with missing rate (Fig. S5). Based on these observations, we compared inter-donor variation between tissue-shared and tissue-specific proteins after stratifying on the reference ion intensity of each protein. Specifically, we defined three strata based on reference ion intensity (log2 scale) as follows: low ($\leq 16$), medium (16-20), and high ($> 20$). The variance of shared and tissue-specific proteins was compared within each stratum. The conclusion is consistent with the unstratified test; the p-values were $1.4 \times 10^{-22}, 1.4 \times 10^{-99}, 2.7 \times 10^{-56}$ in the three strata, respectively (Fig. S6).

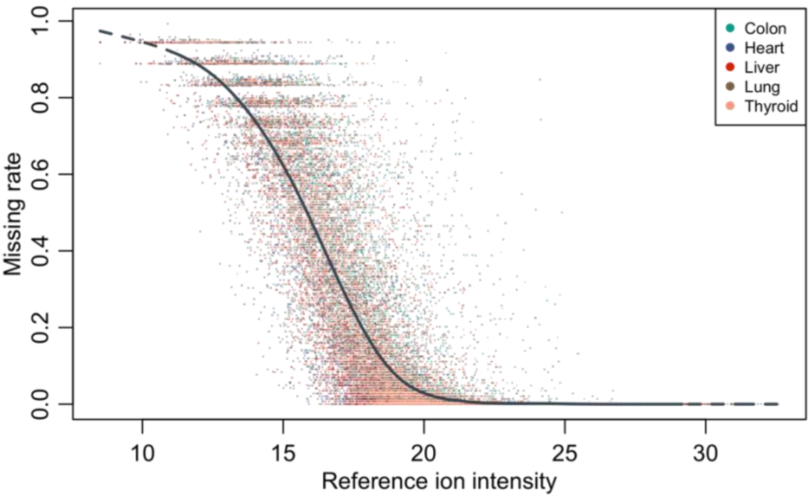


Fig. S5. Reference ion intensity of quantified proteins (x-axis) vs proportions of donors with missing values (y-axis). Solid curve is a smoothing spline combining all tissues.

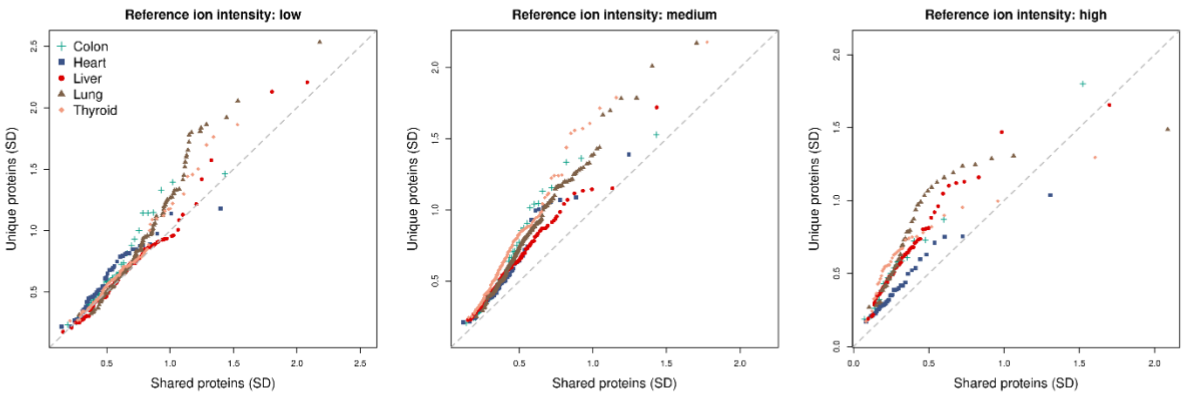


Fig. S6. QQ-plot comparing the variance of proteins that were quantified in all five tissues (x-axis) with proteins that were quantified in a single tissue (y-axis), stratified by reference ion intensity.

#### 3. Factors associated with protein abundance

##### 3.1 Sharing of protein abundance with sex and age across tissues

We used an empirical Bayes hierarchical model, implemented in the R package MASHR, to assess the consistency in sex- and age-association with protein abundance across the five tissues. Effect size and standard error estimates within each tissue were used as input. Only proteins that are tested for sex- or age-association in all five tissues were included. For the analysis of sex-associated proteins, our focus was on the tissue sharing of sex-biased protein abundance at autosomal genes. Therefore, X-linked genes (regardless of observed sex-association) were removed. This led to 5031 proteins assessed for age-association and 4875 proteins assessed for sex-association. The data-driven covariance matrices used in MASHR were set up by applying extreme deconvolution (cov_ed function) on strong signals (proteins that pass q-value<0.1 in any of the tissues)[13]. Other parameters of MASHR used were the factor within which effects are considered to be shared (factor = 0.5, default) and the lfsr threshold for including an effect in the assessment (lfsr_thresh = 0.1).

##### 3.2 Gene set and complex definition

Cytoplasmic ribosome protein complex was defined by HUGO Gene Nomenclature Committee (HGNC) gene group 1054, excluding genes that were designated as

Mitochondrial ribosomal proteins (MRP). Likewise, proteasome complex was defined by gene group 690 (accessed Feb 2024). For comparison of protein-RNA correlation among gene sets presented in Fig. 3B, genes involving absorption, distribution, metabolism, and excretion (ADME) were obtained from Dataset S1 of [14]. For comparing pairwise protein correlations within complexes, we considered all human complexes defined by the CORUM database (Release 3.0 [15]).

##### 3.3 Correlation between protein and mRNA abundance

Pearson correlations between mRNA abundance and protein abundance were calculated for each gene that was measured in at least 72 individuals for protein abundance. Covariates were regressed out from protein and RNA phenotypes using linear regression models, and the residuals were used to compute the correlation. For RNA, the covariates included ischemic time, top five genotype PCs, top 15 PEER factors, sequencing platform and sequencing protocol. For protein, the covariates included ischemic time, top 3 genotype PCs and the same protein PCs as described in the sex- and age-association analysis. The covariate adjustment had little, if any, effects on the overall distribution of the RNA-protein correlations (Fig. S7). Additionally, we assessed the correlation between protein abundance and its corresponding RNA transcript expression. For each gene, we used Ensemble transcript ID (ENST) to match the protein-coding transcript. The distribution of the correlation coeffects was similar.

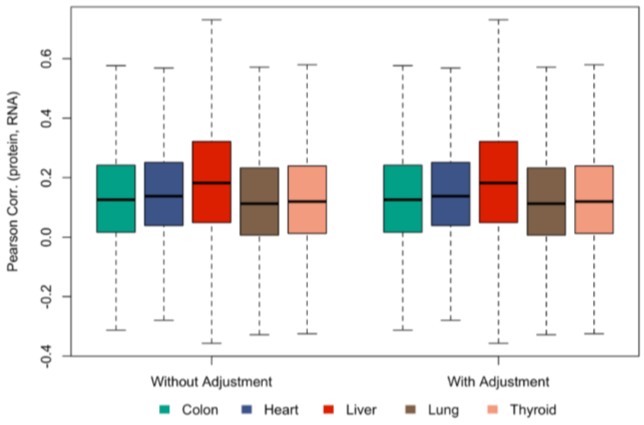


Fig. S7. RNA-protein correlation, with- or without- covariate adjustment.

##### 3.4 Protein-protein correlations in complexes

**Testing correlated protein levels within a complex.** To test the hypothesis that subunits within each complex are more correlated than random set of proteins, we used as test statistics the number of edges that connect pairs of proteins, whose correlations fell below the .5% or above the 99.5% quantiles of all observed pairwise correlations in colon. The null distribution of this edge statistic was computed using a resampling procedure: a pseudo-complex was formed by randomly sampling the same number of proteins as the real complex; the number of edges in the pseudo-complex was counted, using the same, tissue-specific, .5% and 99.5% quantile thresholds. This resampling procedure was repeated 10,000 times. Statistical significance was evaluated by comparing the number of edges in a real complex to those in the pseudo-complexes. The observed edges in ribosome and proteosome were 1473 and 84, respectively, and exceeded the maximum edges observed in 10,000 resampled pseudo-complexes ($p < 10^{-4}$, Fig. S8).

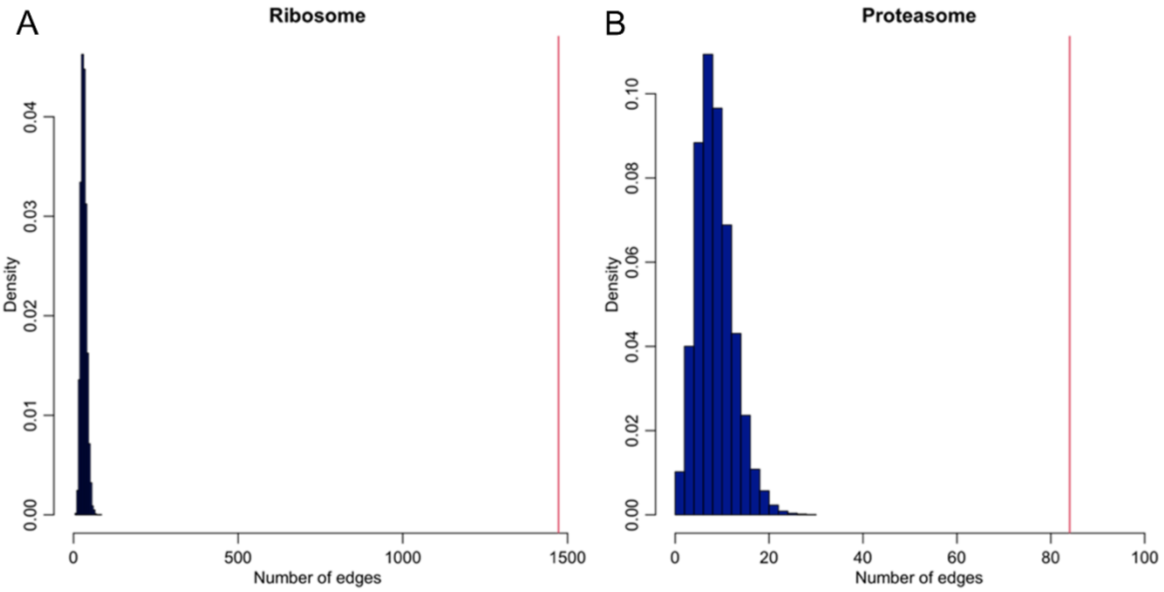


Fig. S8. Network edges based on Pearson correlations between proteins within ribosome (A) and proteasome (B). The red vertical line indicates the observed number of edges; the histogram represents the null distribution of edges in pseudo-complexes of the matching sizes.

**Testing correlated protein levels between ribosome and proteasome complexes.**

We implemented a permutation-based procedure to evaluate whether the correlation pattern *between* the two complexes is more extreme than expected by chance.

Notation: Let $X$ be the complete proteome data matrix, with each row, $X[j,]$, representing a protein and each column, $X[,n]$, a tissue sample. Let $C[j,k] = cor(X[j,], X[k,])$ be the pairwise protein correlation matrix. Define R and P be the submatrix representing the ribosome and proteosome abundance, respectively; that is, R and P are non-overlapping rows of X. We framed the question as testing if correlation structure between members of the two complexes, $v(j,k) = cor(R[j,], P[k,])$, was more extreme than two independently generated complexes.

Test statistic: We defined test statistics using the average quantiles of $v(j,k)$ relative to the upper trianglular elements of $C$, $C^u$. Specifically,

$$\varphi(j,k) = mean(C^u \geq v(j,k)) \text{ and } T(R,P) = mean(-\log(\varphi(j,k))).$$

This observed test statistics, $T(R,P)$ was compared to a null distribution, $\tilde{T}$, which was derived from two levels of permutation. First, we computed a permuted proteome-wide correlation matrix, $\tilde{C}$, by permuting each row (protein) independently. Next, we re-assigned sample ID to ribosome complex proteins by permuted sample labels to generate $\tilde{R}$. This made pairs of proteins from different complexes independent, while preserving the correlation between proteins within each complex. The null distribution of the $T$ statistic was computed on the permuted complexes:

$$\tilde{\varphi}(j,k) = mean(\tilde{C}^u \geq \tilde{v}(j,k)) \text{ and } \tilde{T}(\tilde{R},P) = mean\left(-\log\left(\tilde{\varphi}(j,k)\right)\right).$$

Lastly, the significance of the observed statistic is compared with the permutation null, $p = mean(\tilde{T} \geq T)$.

Applied to five tissues, the p-values for the correlation between ribosome and proteasome were significant in colon ($p = 0.0082$), lung ($p = 0.0050$) and thyroid ($p < 10^{-4}$), but not significant in heart ($p = 0.21$) or liver ($p = 0.4388$).

**4. Protein QTL (pQTL) mapping**

##### 4.1 Phenotype and covariates used in cis-pQTL and trans-pQTL mapping

Both cis- and trans-pQTL mapping was performed in each tissue separately. Autosomal encoded proteins with at least 72 donors with WGS in a tissue were included in pQTL mapping. We used a linear model to assess the association between genotype and quantile-transformed protein phenotypes, adjusting for following covariates: sex, age, ischemic time, the first 3 genotype PCs and the same protein PCs as described in the sex- and age-association analyses. Genetic variants from GTEx V8 were filtered to retain variants with minor allele frequency (MAF) $\geq 0.05$ among individuals with proteomic data; the genotype PCs were recalculated on these individuals.

##### 4.2 cis-pQTL mapping

For cis-pQTL mapping, variants within a window of 1Mb upstream and downstream of the protein coding region were tested. Significance test of cis-pQTL follows a procedure analogous to FastQTL used for GTEx V8 cis-eQTL mapping. We did not apply FastQTL directly because some proteins have non-negligible rate of missing data; treating these observations as 0 or imputing under missing-at-random assumption may lead to inflated Type I error or reduced power. Therefore, we implemented the following procedure. First, for each protein phenotype, a null distribution was derived by permuting the protein phenotypes relative to genetic variants, so that the correlation between variants (linkage disequilibrium, LD) was preserved. The minimum p-value across all variants in the cis-region of a gene was compared to the minimum permuted p-values to obtain the gene-level permutation p-value. The permutation p-value was defined as

$$P_{perm} = \frac{1 + \sum_{n=1}^{N} I(x_n^* \leq x_0)}{1 + N},$$

where N denoted the number of permutations, $x_n^*$ denoted the minimum nominal p-values across all variants in a permutation run, $x_0$ is the corresponding minimum nominal p-value observed in the real data, and $I(\cdot)$ is the indicator function. A pseudo-count of 1 was added to avoid p-values of zero. To reduce the computational burden, a two-stage permutation strategy was used: 1000 permutations were performed for each gene in the first stage; for those genes achieving a permutation p-value $\leq 0.025$, $10^4$ additional permutations were performed, and the final gene-level p-values were calculated from the $1.1 \times 10^4$ permutations. At the proteome level, false discovery rate (FDR) was calculated using gene-level permutation p-values and assuming independence among proteins. Significant cis-pQTLs were defined at FDR<0.1 (Table S5). In subsequent analysis, pGene refers to the protein at a cis-pQTL, while pSNP refers to the index SNP (SNP with the lowest nominal p-value for the pGene).

**4.3 trans-pQTL mapping**

We defined *trans*-region as variants located more than 1Mb up- and downstream of the protein-coding sequence or on a different chromosome. We used the same model, covariates, as well as protein and variants inclusion criteria as for cis-eQTL analysis (described in 4.1). Following the practice of the GTEx trans-eQTL mapping, a protein-level adjusted p-value was calculated by multiplying the smallest nominal p-value (*trans*-pSNP), across all tested SNPs, by $10^6$ and truncated at 1. Lastly, the gene-level adjusted p-values from all protein phenotypes were pooled to compute the tissue-wide FDR via R package, q-value. To investigate if the trans-pSNP impacted the distal protein through a cis-gene, we queried pSNP association in cis-pQTL mapping statistics and used a nominal p-value of 0.01 as a suggestive significance threshold. This led to identifying a putative cis-protein target for 14 of the 30 trans-pQTLs.

As an example, rs2076295 was identified as a trans-pSNP for PKP3 ($p=4.39x10^{-16}$), and a cis-pSNP for DSP ($p=5.54x10^{-27}$) in lung (Fig. S9A-B). Fig. S9C confirms that rs2076295 was not a trans-eQTL of PKP3. Covariation between PKP3 and DSP was observed at both protein and RNA levels. This observed pattern is consistent with a model that PKP3 and DSP are under coordinated regulation at the protein level, which gives rise to the trans-pQTL. The observed, weaker, covariation at the RNA level may reflect feedback mechanisms through which RNA expression is modulated by protein abundance.

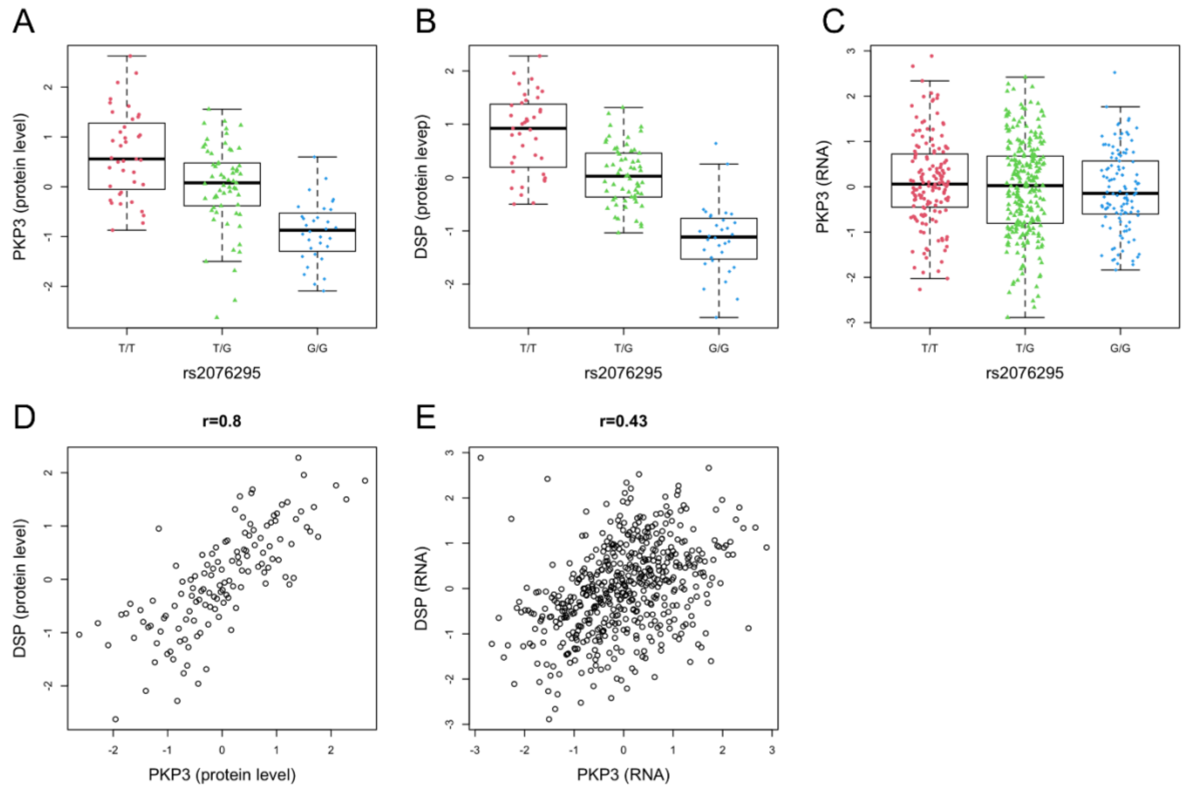


Fig. S9. rs2076295 is a trans-pQTL for PKP3 (A) and a cis-pQTL for DSP (B), but it is not significantly associated with the RNA level of PKP3 (C). DSP and PKP3 shows covariation at both protein (D) and RNA levels (E).

**4.4 Fine-mapping of cis-pQTL with SuSiE**

Within each of the 1981 pQTL loci, we performed fine-mapping analysis using individual-level data and R package susieR[16] with default coverage (0.95) and purity (min_abs_cor=0.5) parameters. For 1356 loci, susieR produced exactly one credible set. Of the remaining loci, 477 and 148 had no credible set and more than one credible set, respectively. At all loci that has at least one credible set, the SNP with the highest posterior inclusion probability (PIP) coincided with the SNP with the largest marginal Z score and 78% coincided with the pSNP. The discrepancy was due to the handling of missing genotypes: susieR imputed the genotype while our computation was restricted to non-missing genotypes. The large fraction of pQTLs with zero or one credible set reflects the challenge in pin-pointing causal variants and independent signals when the sample size is modest. For this reason, subsequent analyses used the pSNP as the proxy for the variant driving the pQTL.

**4.5 Replication of cis-pQTL**

We used two independent, plasma-based, pQTL datasets to evaluate the replication of the cis-pQTL reported here: (1) INTERVAL study, which used an aptamer-based multiplex protein assay (SOMAscan) to quantify 3,622 plasma proteins in 3301 healthy blood donors[17], and (2) the Fenland study, which used a combination of SOMA and Olink arrays to quantify a total of 4,775 proteins in 12,084 participants [9]. We took unique pairs of pSNP-pGene discovered in all five GTEx tissues (n=1835) and queried their corresponding association p-values in INTERVAL and Fenland summary statistics, respectively. The replication was quantified using the $\pi_1$ statistics, which estimated the non-null proportions of GTEx pQTL in each of the replication datasets (Fig. S9). INTERVAL data included 326 unique pSNP-pGene pairs, with $\pi_1 = 0.55$. In Fenland, summary statistics were available for 683 unique pSNP-pGene pairs, with $\pi_1 = 0.67$. Furthermore, we observed that the estimated allelic effects in Fenland are highly consistent with the corresponding allelic effects in GTEx, regardless of whether the association is statistically significant (Extended Data Fig. 7).

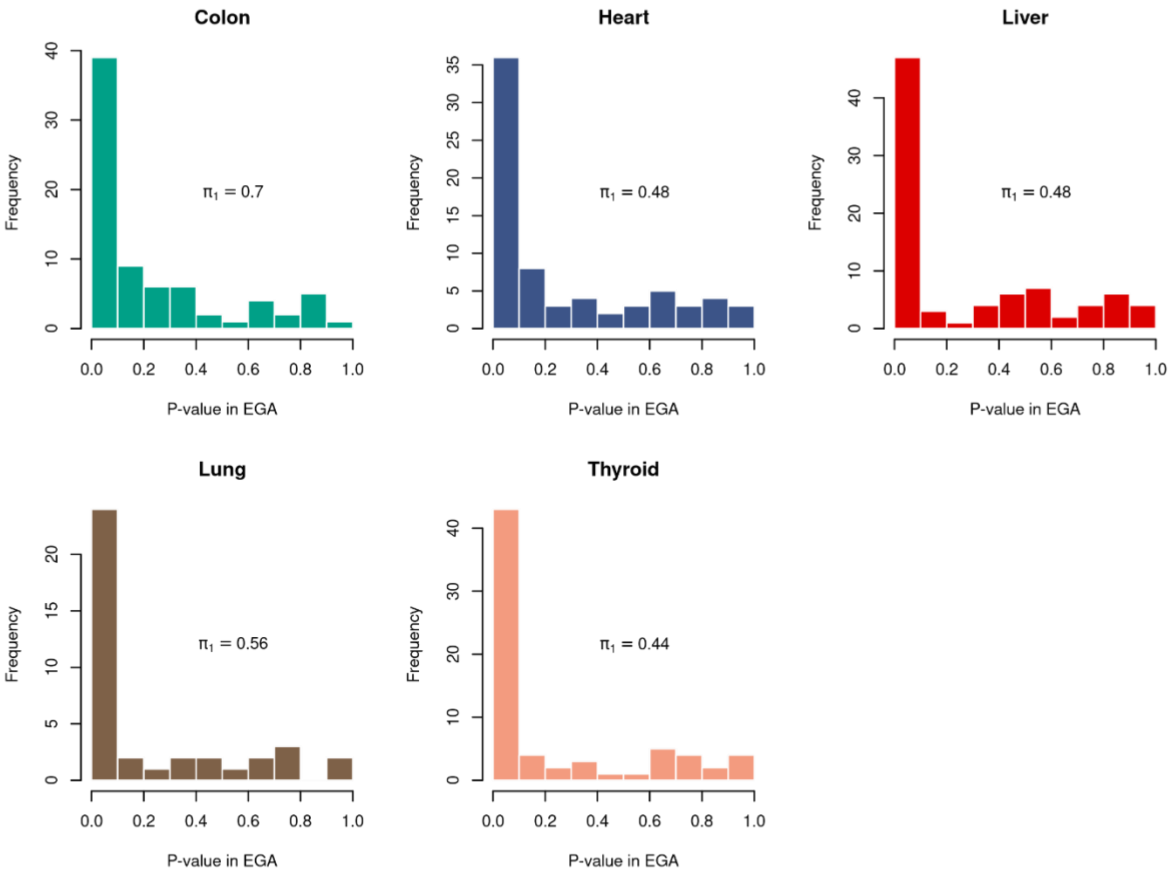


Fig. S10. Replication of cis-pQTL in INTERVAL. For pQTLs discovered in each of the five GTEx tissue, replication in INTERVAL was quantified by the $\pi_1$ statistic. Replication in Fenland and the consistency in effect sizes are presented in Extended Data Fig. 7.

#### 4.6 Tissue-sharing of cis-pQTL

We used two complementary approaches to assess the degree of pQTL sharing across tissues. First, taking a pair of tissues, we treated one tissue as the focal tissue to define pSNP-pGene pairs. The corresponding p-values and the allelic effects were queried in the second tissue. We quantify the degree of tissue-sharing using the FDR and $\pi_1$ statistics, computed by R package qvalue (Fig. S11). The correlation of allelic effects between pairs of tissues are shown in Extended Data Fig. 6.

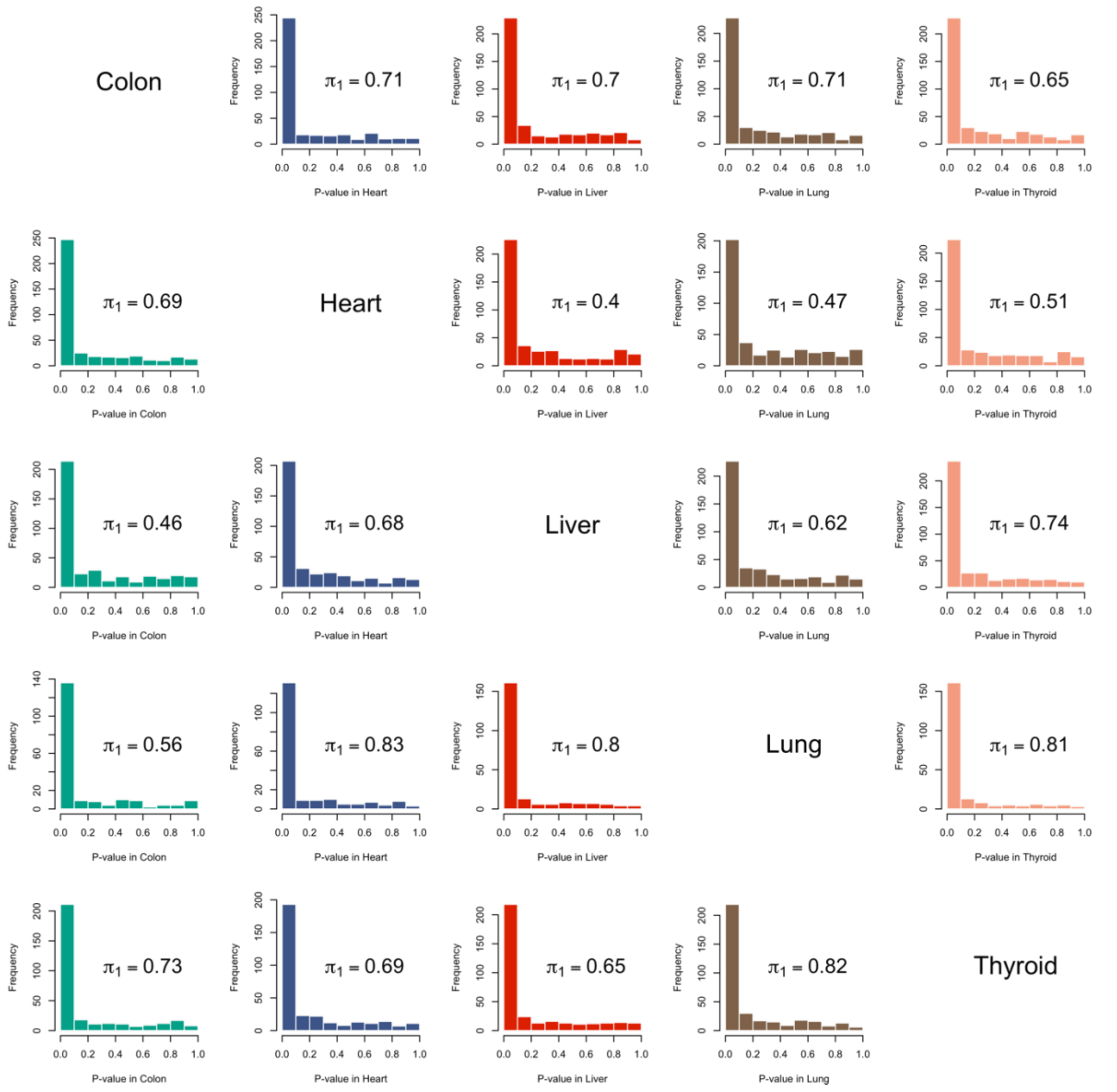


Fig. S11. Tissue-sharing of cis-pQTL, quantified by replication proportion, $\pi_1$. pSNP-pGene pairs are defined in one tissue (focal tissue, x-axis); the corresponding p-values from a second tissue (y-axis).

In a second approach, we used MetaSoft, a Bayesian meta-analysis approach that combined pSNP-pGene association statistics across all five tissues[18]. MetaSoft produces a posterior estimate on the number of tissues in which a pQTL is likely active. For this analysis we included pSNP-pGene pairs that were deemed significant in at least one of the five tissues (Extended Data Fig. 6A)

#### 5. Genomic and transcriptomic context of cis-pQTLs

##### 5.1 Functional annotation of cis-pQTL

To facilitate the comparison of the annotation of pSNP with that of eSNP (Fig. 4C), we used the variant effect prediction (VEP) data generated in GTEx V8 to annotate pSNPs for the following categories: enhancer, promoter, open chromatin, promoter flanking, CTCF binding, 3’ UTR, 5’ UTR, frame shift, intron, missense, non-coding transcript exon, splice acceptor, splice donor, splice region, stop gained, and synonymous. We note that this annotation considered the impact of a variant on *all* proximal genes, and it was possible for a variant to be included in multiple categories. pSNPs across all tissues were pooled and the relative proportions for the annotation category were compared to the background of all genetic variants (passing MAF of 0.01 in GTEx v8). To characterize the proximity between pSNP-pGene, we computed the distance between pSNP and the transcription start sites (TSS) of the pGene, for pSNPs that were not located within the coding regions.

##### 5.2 Gene-specific functional annotation

We ran Ensembl’s Variant Effect Predictor (VEP v88, GENCODE v26) to identify putative protein altering variants (PAV) in a gene-specific manner. Variant consequence annotations in the following categories are included: missense_variant, splice_donor_variant, splice_acceptor_variant, stop_gained, start_lost, stop_retained_variant, inframe_insertion, inframe_deletion and frameshift_variant.

##### 5.3 eQTL-mediated pQTL

To characterize the overlap in regulation at the RNA level and at the protein level, we used two complementary approaches. First, we performed a mediation analysis[19] to formally test the hypothesis that the pSNP-protein association was mediated through regulation at the RNA expression level. At each pQTL, the mediation analysis compared two linear models, both with the protein abundance as the outcome: one includes, as a covariate, the predicted RNA abundance, R, and the other without (model 0):

**Model 0**. $\text{Protein} = \mathrm{b}_0 + \mathrm{b}_1\,\mathrm{G} + \sum b_j Z_j + \varepsilon_0$,

**Model 1**. $\text{Protein} = \mathrm{a}_0 + \mathrm{a}_1\,\mathrm{G} + \alpha R + \sum a_j Z_j + \varepsilon_1$,

where G denoted the genotype at the pSNP, R denoted the predicted RNA and Z's were additional covariates, which included sex, age, ischemic time, protein PCs (same as used in the pQTL analysis), and the top three genotype PC. The predicted RNA was computed by a linear model similar to that used in the eQTL analyses, and included as predictors, genotype of the pSNP, age, sex, ischemic time, genotype PCs and top 15 PEER factors. The statistical significance of the mediation effect at each pQTL was determined using a bootstrap method for testing the null hypothesis, $\mathrm{H}_0\!: \mathrm{a}_1 = \mathrm{b}_1$ against the two-tailed alternative hypothesis: $\mathrm{H}_1\!: \mathrm{a}_1 \neq \mathrm{b}_1$. Multiple comparison was accounted for by computing q-values across all pQTLs.

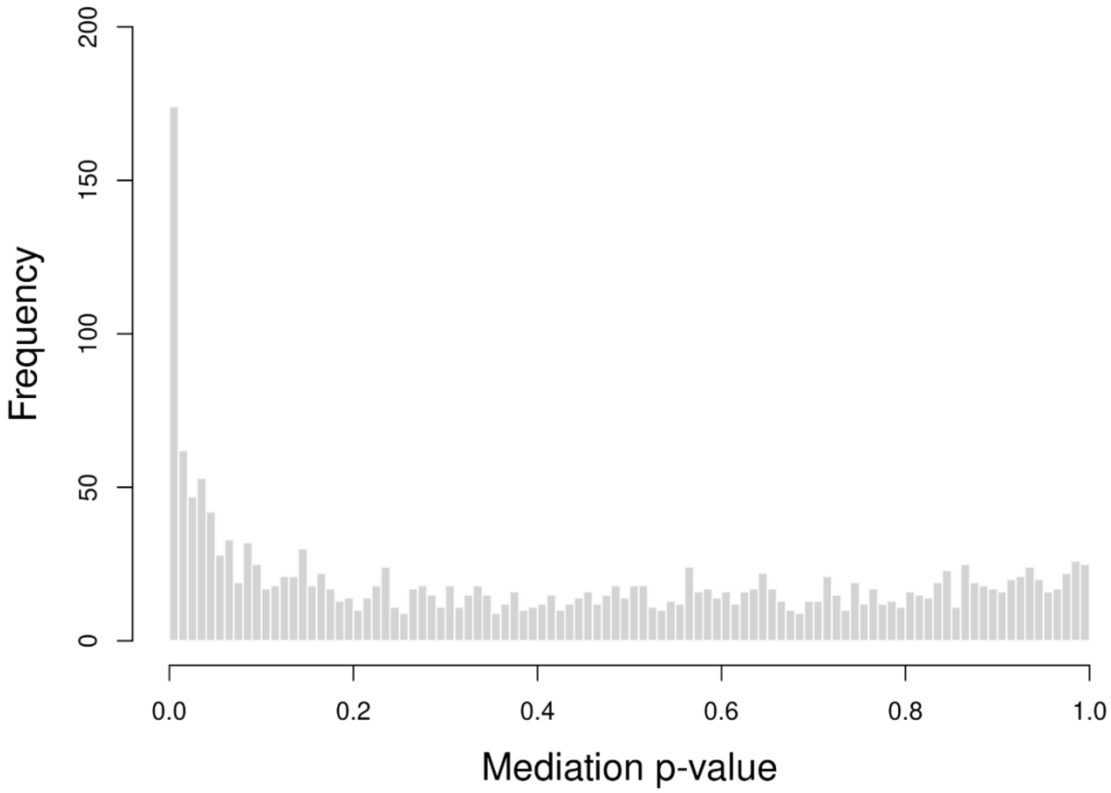


Fig. S12. Results of mediation analysis: histogram of p-value.

##### 5.4 Defining eQTL-supported cis-pQTLs

Since mediation analysis may be underpowered, we also used a second, and more liberal, criterion to assess RNA-level support for pQTL. We extracted the SNP-mRNA association information from the GTEx (V8) eQTL summary statistics in the matching tissue. We define a pQTL as *eQTL-supported* if the pSNP was associated with RNA abundance in the matching tissue, using significance threshold set in the GTEx cis-eQTL analysis (Supplemental Method 4.5 of [20]) and had consistent direction of allelic effect. By this criterion, 49% of pQTL had eQTL-level support. The correlation of allelic effect on mRNA and protein was computed for each tissue and overall pSNPs, regardless of if the SNP-mRNA association reached cis-eQTL significance.

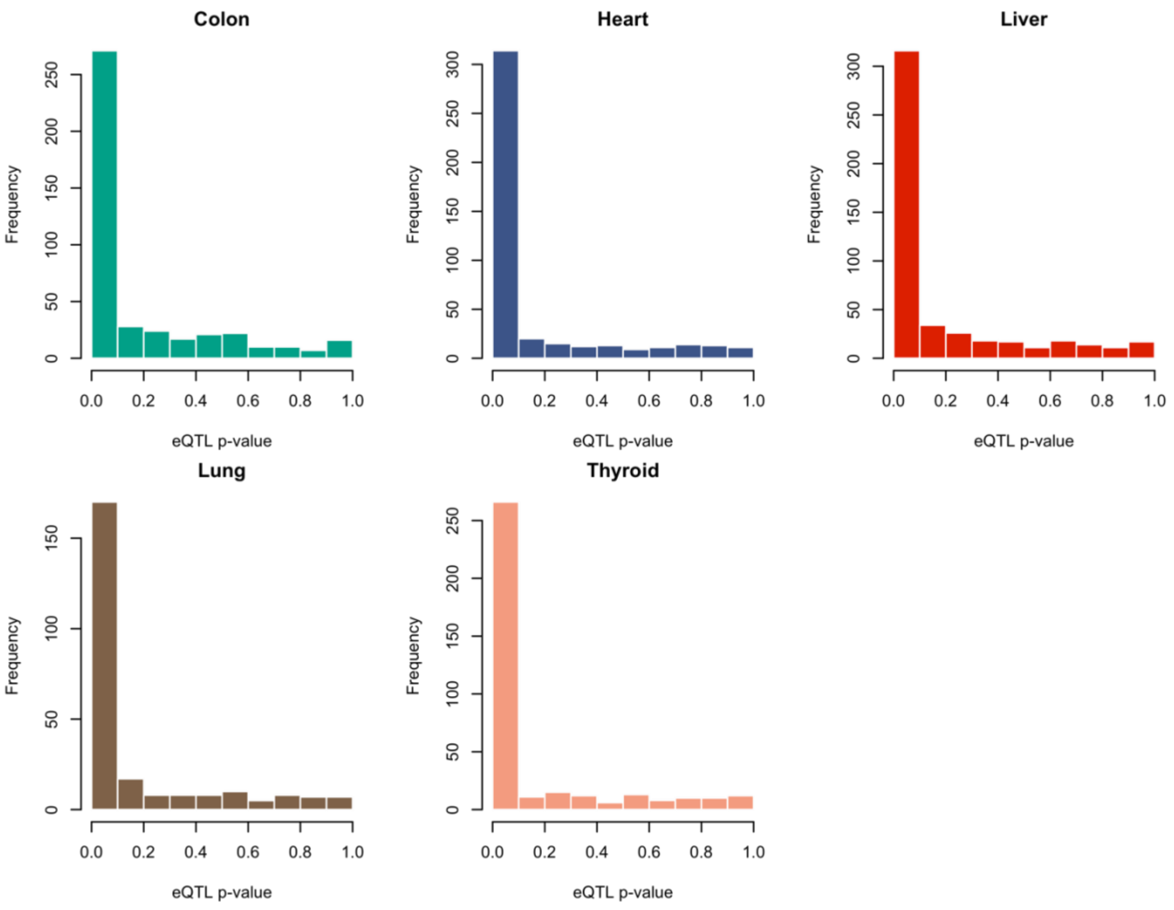


Fig. S13. Transcriptomic support at pQTLs. Distribution of eQTL p-values for SNP-gene pairs corresponding to significant pQTL.

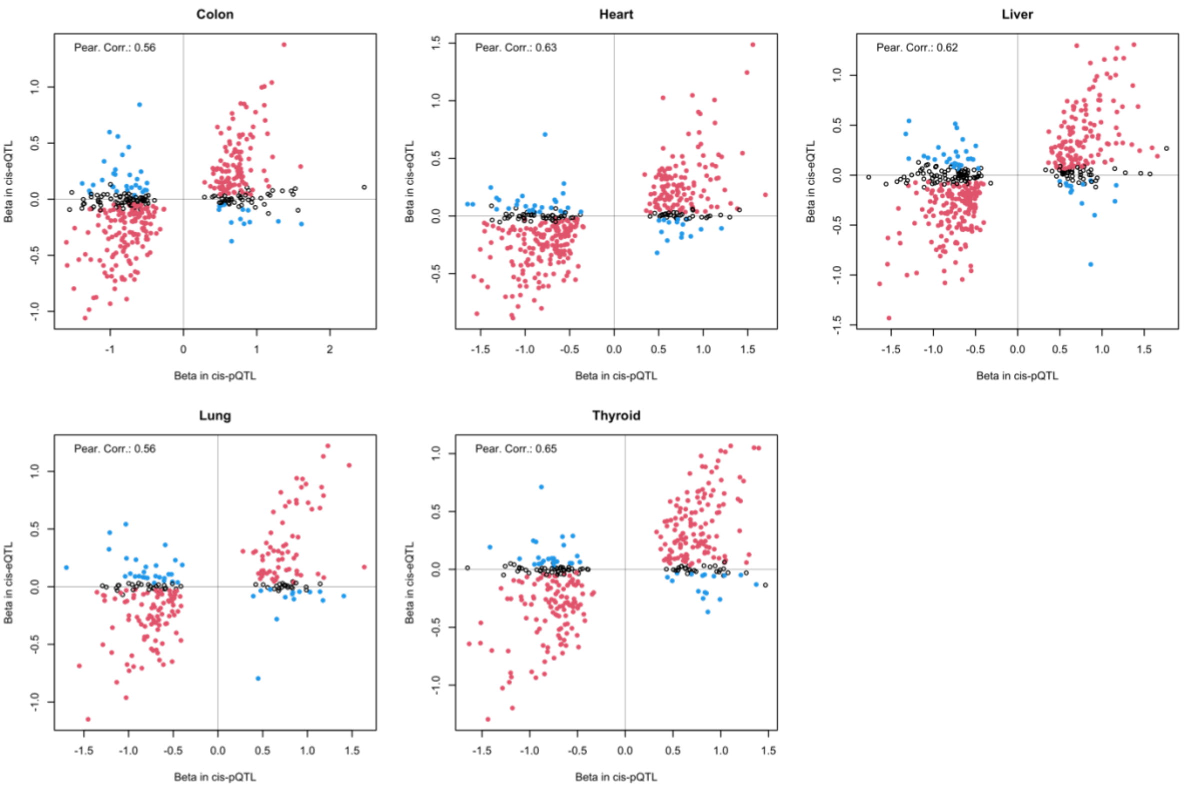


Fig. S14. Comparison of allelic effects in cis-eQTL and cis-pQTL for pSNP-pGene pairs in each tissue.

As expected, pQTLs supported by eQTLs exhibited elevated RNA-protein correlations compared to genes without pQTL or a pGene not supported by an eQTL.

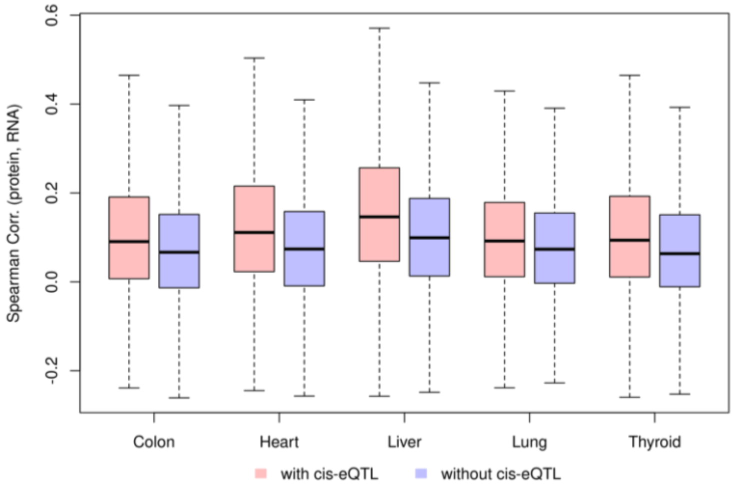


Fig. S15. RNA-protein correlations at genes with eQTL-supported pQTLs (pink) and at genes without pQTLs or with pQTLs but no eQTL support (blue).

**5.5 Transcript-level QTL (trQTL) mapping**

Because the primary goal of this analysis was to estimate the proportions of pQTLs that were likely driven by regulation through transcript-level RNA variation, we only tested the association between a pSNP and its corresponding protein-coding transcript abundance, provided that the transcript abundance was quantified by GTEx analyses. A small number of pGenes, whose protein was translated from multiple transcript isoforms or whose transcript data was missing, were excluded. Statistical significance for trQTL was determined using the gene-level eQTL p-value threshold established for each gene in GTEx V8.

**5.6 Additional information of MIF example**

In examining the gene-level and transcript-level evidence at pQTLs, we noted discordant examples that arise from genomic features that complicate gene- and/or transcript-level quantification using short-reads data. An example illustrating this latter case was found at a liver pQTL at gene *MIF* (Extended Data Fig. 4). The reference allele of the pSNP, rs2739338-T, was associated with higher protein abundance ($p < 9.2 \times 10^{-18}$). The association was replicated in the much larger plasma-based Fenland cohort with the consistent direction of allelic effect ($p < 3.11 \times 10^{-9}$, [9]). Surprisingly, the rs2739338-T was associated with decreased gene-level RNA abundance ($p = 3.6 \times 10^{-29}$) and no significant sQTL was identified in *MIF* in any of the 49 GTEx tissues on the basis of intron excision ratios estimated from short-reads RNA-seq data[20]. Leveraging a newly generated long-reads RNA sequencing dataset that quantified *MIF* transcript abundance in a small subset of GTEx tissue-samples, we confirmed the accuracy of each *MIF* transcript quantification constructed using the short-reads RNA-seq data (Extended Data Fig. 4D)[21]. *MIF* encodes three known alternative transcripts, of which only one (ENST00000215754) leads to protein product. The association between rs2739338 genotype and ENST00000215754 abundance was evaluated using all liver tissues (N=208). This analysis confirmed that rs2739338-T was significantly associated with an increased level of the protein-coding transcript ($p = 5.18 \times 10^{-32}$), consistent with the pQTL direction and opposite to the gene-level eQTL direction. Indeed, across 144 liver tissues, the ENST00000215754 transcript level was inversely correlated with the gene-level abundance. More surprisingly, each of the three MIF transcripts was inversely correlated with the gene-level RNA level. In this case, the discrepancy between gene-level eQTL and trQTL/pQTL likely arose from an overlapping anti-sense transcript, MIF-AS1, which was erroneously included in the quantification of gene-level MIF RNA abundance but not included in the transcript quantification. This example underscores the importance of precise gene- and transcript-level quantification for interpreting transcriptional consequences, which can be achieved through the advances in sequencing technology and analytic strategies.

#### 7. Additional analyses of NAT2

We performed peptide- and haplotype-based analyses to eliminate the possibility that the pQTL of *NAT2* in liver is due to detection bias of non-reference peptide variants. Six protein-coding variants used to define *NAT2* alleles by the Pharmacogene Variation Consortium ([https://www.pharmvar.org/gene/NAT2](https://www.pharmvar.org/gene/NAT2)) have a minor allele frequency greater than 0.01 in 1000Genome phase 3 data (European populations) and passed the 1% MAF filter in GTEx.

| variants | Ref/Alt | Ref_AF | AminoAcids | Note |
|---|---|---|---|---|
| rs1041983 | C282T | C=0.6948 | Y94Y | synonymous |
| rs1801280 | T341C | T=0.5507 | I114T | Non-synonymous |
| rs1799929 | C481T | C=0.5646 | L161L | synonymous |
| rs1799930 | G590A | G=0.7177 | R197Q | Non-synonymous |
| rs1208 | G803A | A=0.5616 | R268K | Non-synonymous |
| rs1799931 | G857A | G=0.9771 | G286E | Non-synonymous |
| rs4921913 | C>T | C=0.2435 | | pSNP |

*Ref/Alt: Reference and alternate DNA variants.

*Ref_AF: Reference allele frequency in the 1000 Genomes European sub-group.

Fig. S16. Summary of variants in NAT2. Of variants used by the Pharmacogene Variation Consortium to define NAT2-alleles, six coding variants exceeded allele frequency of 1%. The last variant is the pSNP.

NAT2 haplotypes, defined by the six coding SNPs and the non-coding pSNP (rs4921913) were extracted using the GTEx (V7) phased genotype data. Nine unique haplotypes were observed in the liver tissues analyzed in this study. The haplotype and the corresponding NAT2 allele names are tabulated below.

| haplotype | N | Allele | Haplo-group |
|---|---|---|---|
| 0110001 | 82 | *5B | B |
| 1000100 | 4 | *13A | A |
| 0000100 | 36 | *4 | A |
| 1001101 | 58 | *6A | C |
| 0100001 | 1 | *5C | B |
| 0110101 | 8 | *5A | C |
| 1000111 | 5 | *7B | C |
| 0000101 | 1 | *4 | A |
| 0000000 | 3 | *12A | A |

Fig. S17. Haplotype frequencies in GTEx liver donors. Each bit in the haplotype string corresponds to a variant in Fig. S16 in the same order, with 0 representing the reference allele and 1 for derived allele. As rs1208 overlaps a peptide spectrum used for quantifying protein abundance, highlighted haplotypes carry a PAV. Haplogroup was assigned based on the estimated haplotype effects and SNP alleles.

NAT2 protein quantification in the present study was based on the spectra of three peptide fragments: EQYITNK, DNTDLVEFK, and TLTEEEVEEVLR. The former two peptides did not overlap any PAV. The third peptide overlaps with rs1208, (R268K), for which the reference variant included in the peptide search database corresponded to the allele *NAT2*5B*, **5C* and **12A*. We estimated the haplotype association with each of the three peptides using a linear regression model, in which the response presented peptide level abundance ratio or protein abundance ratio, and haplotype dosage (0,1,2) were used as predictors. *NAT2*5B* and *NAT2*5C* were combined and used as the reference in the regression because they were the most frequent haplotypes, and their amino acid sequence matched the reference alleles in the search database on all three peptide spectra. For this analysis, peptide abundance ratio was not normalized and replicates between samples were not combined; protein-level abundance was normalized for sample and run effects, but replicates were not combined. The estimated haplotype effects are shown in Extended Data Fig 8.

With respect to the two peptides that did not carry PAV, the three “rapid” acetylator alleles (*NAT2*4, *12* and **13*) were associated with increased abundance compared to *NAT2*5BC,* while the “slow” acetylator alleles (*NAT2*5A, *6A*, and **7B*) shows similar abundance as *NAT2*5B*, consistent with the pattern of association at the protein-level. With respect to the peptide carrying the missense variant rs1208, all *NAT2* alleles, except **12A* (carrying the reference allele of rs1208), showed decreased abundance compared to *NAT2*5B,* consistent with a biased measurement of non-reference peptide variants. However, if the pQTL had arisen due to such bias, one would expect a decreased protein abundance associated with *NAT2*4*. The opposite was observed: the pSNP rs4921913-C allele was linked to *NAT2*4,*12A* and **13A*, and was associated with an increased protein abundance. Therefore, the association could not be attributed to the reduced quantification of non-reference peptide variants. To summarize the relationship between diplotype and protein abundance, we combined haplotypes into groups based on the estimated haplotype effects and shared genetic variants (Fig 5; Extended Data Fig 8) . Specifically, “rapid” acetylators, *NAT2*4, *12* and **13* were combined as labelled as [A], *NAT2*5A, *6A,* and **7B* were combined and labelled as [C]. *NAT2*5C* (N=1) was grouped with **5B* (N=82) and used as the reference group [B].

**7. Gene ontology (GO) enrichment analysis**

All GO enrichment analysis were performed using Bioconductor package fgsea (v1.22.0) and the GO database of MSigDB v7.4[22]. The analyses of sex- and age-associated proteins applied the classic gene set enrichment analysis (implemented in function fgsea) to the regression z-score with signs, so that enrichment at both extremes – male and female, old and young – could be identified. The enrichment analysis of sex-associated proteins was restricted to proteins coded by autosomal genes, as many sex-associated proteins represented escape of X-inactivation. All other enrichment analyses used the over-representation function, fora, which was based on a hypergeometric test contrasting a foreground gene set with a background (reference) set. The background gene sets depended on specific analysis and are listed in Table S7 (below). For all enrichment analyses, an adjusted p-values of 0.05 was used as the significance threshold; the results are provided in Table S3.

Table S7 Foreground and background in GO enrichment analysis

| Gene-set | Foreground | Background | Test in each tissue or across all tissues |
|---|---|---|---|
| Tissue-shared | Proteins quantified in all five tissues | Proteins quantified in at least one of the five tissues | Across all tissues |
| Tissue-specific | Unique genes in each tissue | Proteins with non-missing observation number >= 1 in each tissue | Each tissue |
| Most variable | Top 5% most variable proteins in each tissue (require N>20) | Proteins with non-missing observation number N>20 | Each tissue |

- For enrichment of sex-associated proteins, X-linked proteins are removed from the analysis as many of the sex-associated proteins represented escape from X-inactivation.

**Web Resources**

GTEx donor characteristics and whole genome sequencing data (WGS) are available on the database of Genotypes and Phenotypes (dbGaP) (accession no. phs0000424.v8).

RNA quantification and single-tissue eQTL summary statistics for GTEx v8 are available on the GTEx Portal (https://www.gtexportal.org/home).

Protein-coding transcript translation sequences: GENCODE v26 (https://ftp.ebi.ac.uk/pub/databases/gencode/Gencode_human/release_26/gencode.v26.pc_translations.fa.gz).

Protein complex: CORUM database v3.0 (https://mips.helmholtz-muenchen.de/corum/) and STRING database v12.0 (https://string-db.org/).

GWAS results: GWAS catalog (https://www.ebi.ac.uk/gwas/, downloaded on 2024-01-18) and Phenoscanner v1.0 (http://www.phenoscanner.medschl.cam.ac.uk/, downloaded on 2024-01-18).

Variants effects prediction (VEP): https://www.ensembl.org/info/docs/tools/vep/index.html.

Locus zoom (http://locuszoom.org/, visited 2024-01-24).
